# Identifying priority populations for HIV interventions using acquisition and transmission indicators: a combined analysis of 15 mathematical models from 10 African countries

**DOI:** 10.1101/2025.03.17.25324139

**Authors:** Romain Silhol, Ross D. Booton, Kate M. Mitchell, James Stannah, Oliver Stevens, Dobromir Dimitrov, Anna Bershteyn, Leigh F. Johnson, Sherrie L. Kelly, Hae-Young Kim, Mathieu Maheu-Giroux, Rowan Martin-Hughes, Sharmistha Mishra, Jack Stone, Robyn Stuart, John Stover, Peter Vickerman, David P. Wilson, Stefan Baral, Deborah Donnell, Jeffrey W. Imai-Eaton, Marie-Claude Boily

**Affiliations:** MRC Centre for Global Infectious Disease Analysis, School of Public Health, Imperial College London, London, UK; HIV Prevention Trials Network Modelling Centre, Imperial College London, London, UK; Department of Nursing and Community Health, Glasgow Caledonian University, London, UK; Department of Epidemiology and Biostatistics, School of Population and Global Health, Faculty of Medicine and Health Sciences, McGill University, Montréal (QC), Canada; Vaccine and Infectious Disease Division, Fred Hutchinson Cancer Center, Seattle, WA, USA; Department of Population Health, New York University Grossman School of Medicine, New York, NY, USA; Centre for Integrated Data and Epidemiological Research, School of Public Health, University of Cape Town, Cape Town, South Africa; Burnet Institute, Melbourne, VIC, Australia; Department of Medicine, University of Toronto, Toronto, ON, Canada; Population Health Sciences, Bristol Medical School, University of Bristol, Bristol, UK; Avenir Health, Glastonbury, CT, USA; Bill & Melinda Gates Foundation, Seattle, WA, USA; Johns Hopkins University, Baltimore, MD, USA; Center for Communicable Disease Dynamics, Department of Epidemiology, Harvard T. H. Chan School of Public Health, Boston, MA, USA

**Keywords:** AIDS, HIV, HIV incidence, key populations, female sex workers, men who have sex with men, sub-Saharan Africa

## Abstract

**Background:** Characterising disparities in HIV infection across populations by gender, age, and HIV risk is key information to guide intervention priorities. We compared 9 models representing 15 different settings across Africa to assess how indicators measuring HIV acquisitions, transmissions, or potential long-term infections influence estimates of the contribution of different populations to new infections, including key populations (KPs, including female sex workers (FSW), their clients, men who have sex with men).

**Methods:** We evaluated four indicators: I_1_) *acquisition indicator* measuring the annual fraction of all new infections acquired by a specific population, I_2_) *direct transmission indicator* measuring the annual fraction of all new infections directly transmitted by a specific population, I_3_) 1-year and I_4_) 10-year transmission population-attributable fractions (*tPAFs*). tPAFs measure the fraction of new infections averted if transmission involving a specific population was blocked over a specific time period. We compared estimates of the four indicators across 7 populations and 15 settings and assessed if the contribution of specific populations is ranked differently across indicators for 10 settings.

**Findings:** Indicators identified distinct priority populations as the largest contributors: The *acquisition indicator* (I_1_) identified women aged 25+ years outside KPs as contributing the most to acquired infections in 8/10 settings in 2020, but to direct transmissions (I_2_) in only two settings. In 6/10 settings, the 10-year *tPAFs* (I_4_) identified non-KP men aged 25+ years and clients of FSW as the largest contributors to HIV transmission. Notably, non-KP women aged 15-24 years acquired (I_1_) more infections in 2020 (median of 1·7-fold across models) than they directly transmitted (I_2_), while non-KP men aged 25+ years and clients of FSWs transmitted more infections than they acquired in all but one model (median: 1·4 and 1·6-fold, respectively). Estimates of the 10-year *tPAFs* accounting for transmission in the long-term were substantially larger than the *direct transmission* indicator for all populations, especially for FSW (median: 2·0-fold).

**Interpretation:** Indicators that reflect HIV acquisitions and transmissions over the short and long term can be utilised to capture the complexity of HIV epidemics across different populations and timeframes. The added nuance would improve the effectiveness of the HIV prevention response across all populations at risk.

**Funding:** NIH, MRC.

**Research in context:** *Evidence before this study:* Measures of the distribution of HIV acquisition across population groups are commonly used for assessing the contribution of populations to new HIV infections and prevention priorities. However, alternative indicators documented in the literature reflect transmissions or potential long-term effects. It is unclear how the choice of indicator affects the identification of populations that require additional prevention and treatment efforts to accelerate progress towards ending AIDS. We searched PubMed on March 08, 2025, with the terms (HIV) AND (Africa*) AND (acqui*) AND (transm*) AND (model*), with no language or publication date restriction, and identified no meta-assessment or mathematical model comparison studying differences in estimates of the fraction of all infections acquired and transmitted by a population when using different epidemiological indicators.

*Added value of this study:* Using estimates from 9 models representing 15 different epidemic settings across Africa, we studied indicators of HIV epidemic contribution for 7 populations, including female sex workers, their clients, men who have sex with men, and non-key populations stratified by gender and age. We measured four commonly reported indicators of HIV contribution. One focused on acquired infections and the other three focused on transmissions. We found that estimates from these different indicators can differ greatly for the same model and population, to the extent that they identify different populations for prioritising interventions to accelerate HIV incidence declines. The acquisition-focused indicator (i.e. fraction of all infections acquired by a given population), the most used and communicated by UNAIDS, substantially underestimates the large contribution of men, and particularly male clients of female sex workers, to ongoing HIV transmission.

*Implications of all the available evidence:* The choice of indicators measuring a population’s contribution to the HIV epidemic should be carefully considered and precisely defined. Modelling teams working in partnership with government, implementers, funders, and community members should systematically report both acquisition- and (long-term) transmission-focused indicators, instead of only measuring acquisitions in the short term as currently done. Multiple indicators will more comprehensively capture the potential impact of prevention efforts addressing acquisition and transmission risks of different vulnerable populations.

## Introduction

Identifying the populations that most need additional HIV prevention and treatment efforts can help accelerate the overall reduction in HIV incidence and reach the goal of ending AIDS by 2030^1–3^. The “Know your epidemic, know your response” approach endorsed by the Joint United Nations Programme on HIV/AIDS (UNAIDS) first necessitates an understanding of how current prevention and treatment gaps among different populations influence their contribution to new HIV acquisitions and transmissions^1,4^.

The annual fraction of new HIV infections acquired across population groups (“*acquisition indicator*”) is commonly used to prioritise populations with greatest prevention needs^5^. Estimates of this indicator are easy to interpret, reported by UNAIDS and frequently considered in national response plannings^5–9^. Although the *acquisition indicator* reflects the state of the HIV epidemic and current gaps in HIV prevention, prior work has shown that it only reflects one aspect of the potential overall contribution of each population to HIV incidence, potentially misidentifying priority populations. This is because 1) it focuses on HIV acquisitions among each population and does not reflect the potential contribution of people living with HIV (PLHIV) to HIV transmissions, and 2) it underestimates the longer-term contribution of populations to new infections through onward transmissions from their partners (indirect transmissions)^10,11^. In particular, the *acquisition indicator* may underrepresent the epidemiologic importance and population-wide impacts of addressing HIV service needs of key populations (KPs), such as female sex workers (FSW) or gay, bisexual, and other men who have sex with men (MSM) who have small population sizes^11–13^.

The *direct transmission* indicator measures the annual fraction of new HIV infections directly transmitted by a specific population. However, this has seldom been used despite being easy to interpret, because it is harder to measure or relate to empirical data than the *acquisition indicator*^14,15^. In recent years, the transmission population-attributable fraction (*tPAF*) has emerged as an alternative to the *direct transmission* indicator by measuring the fraction of all new infections that could be averted over a fixed time-period by preventing all transmission from specific populations. The tPAF has the advantage of adequately measuring the long-term contribution of unmet HIV prevention and treatment needs among different populations to new infections as it can reflect both direct and indirect transmission^16,17^. However, the *tPAF* is almost never directly comparable to empirical data (far less than the *acquisition indicator*), its calculation requires a dynamic transmission model, and it is also more difficult to interpret and communicate^17^.

Different indicators (especially *acquisition* vs *transmission indicators*) may identify different priority populations (i.e., populations contributing the most to new HIV infections) in a specific setting. This is because their estimates differently reflect differences in contact patterns, biological factors modulating HIV susceptibility and/or transmissibility (e.g. sexually transmitted infection, increased HIV susceptibility of young women), and differences in epidemic contexts as measured by HIV prevalence and levels and types of intervention already in place^11,18,19^.

Currently only a few mathematical modelling studies have reported estimates using different indicators and even fewer have examined their dissimilarities and influence on the identification of priority populations for HIV prevention^11,20–23^. These studies often highlighted that *acquisition indicators* underestimate the potential impact of addressing the prevention needs of KPs to reduce overall HIV incidence^11,20^. This underscores the need to understand what the most appropriate indicators are for informing the optimisation of future interventions for different populations, based on the intervention effect (i.e., reducing acquisition or transmission risks, or both)^4,11^.

The objectives of this study were to conduct a mathematical model comparison analysis of HIV transmission in highly burdened African countries to examine whether the different indicators identify the same priority populations contributing most to new infections, and to what extent and in which contexts (e.g. indicator type, region, population of interest) indicators diverge the most.

## Methods

### Models’ inclusion criteria and selected populations

We requested outputs from mathematical modelling teams that regularly publish on HIV and are part of the HIV Modelling Consortium^24^. To be included in the analysis, mathematical models had to be calibrated to demographic and HIV epidemiological data from an African setting, include FSWs and their clients (CFSW) and non-key populations (non-KPs), and provide outputs for at least one of the following 3 KPs: 1) FSW, 2) CFSWs, and 3) MSM, with the suggestion to provide additional estimates for non-KP women aged 4) 15-24 years old and 5) 25+ years old, and non-KP men aged 6) 15-24 years old and 7) 25+ years old.

Modelling teams provided details on key model and parameter assumptions, including KP sizes, ART coverage and viral load suppression over time. No specific restrictions were specified for model structure, parameters and calibration data. Baseline scenarios assumed existing levels of intervention (e.g., HIV testing rates). Modelling teams were requested to produce estimates for four HIV contribution indicators for the years 2010 and 2020 for seven defined populations.

### HIV contribution indicators considered

We considered four different HIV contribution indicators (further described in appendix pp 8-9). The a*cquisition indicator (I_1_)* estimates the fraction of all newly acquired infections in one year that are among each population. *Direct transmission indicator (I_2_)* is the fraction of new infections in one year that are directly transmitted by each population. The 1-year and 10-years *transmission population attributable fractions (tPAF)* measure the fraction of all new infections which could be averted by blocking all transmissions from a population (but not their risk of acquiring HIV) over 1 year (*I_3_*) and 10 years (*I_4_*)^11,17^. Since blocking all transmissions from a specific population may also avert future transmissions from their partners, the *tPAF* reflects the long-term impact of hypothetically preventing all transmission from a population^17^. The sum of the *tPAF* across distinct groups can exceed 100% as individuals can acquire HIV from different populations.

### Models included in the analysis

We included estimates from 15 modelled epidemic settings: 9 different transmission dynamic models calibrated to 10 settings in Eastern and Southern Africa (ESA)^21,25–29^ and 5 settings in Western and Central Africa (WCA)^20,22,23^ (Table S1).

Nine of the 15 sets of model estimates (henceforth referred to as “models”) reported estimates for all non-KP and FSW and their clients, using all indicators (Table 1)^20,25,27^. The Maheu-Giroux model provided estimates for each population but only for indicators I_1_, I_3_ and I_4_^23^. Two models (*Stone* and *Silhol Cameroon*) were not age-structured and therefore could not provide estimates for age-stratified non-KP groups^21,22^, while three other models (*Goals*, *Thembisa*, and *Mishra*) did not report estimates for all non-KP groups^26,28,29^. HIV pre-exposure prophylaxis (PrEP) was only incorporated in the *Thembisa* model^28^.

**Table 1.** Characteristics of models, populations, and indicators used for the analysis.

|  |  |  |  |  |  |  |  |  |  |
| --- | --- | --- | --- | --- | --- | --- | --- | --- | --- |
| Model | Country | Populations included and reported on |  |  |  |  |  |  | Indicators reported |
|  |  | Younger non-KP women | Older non-KP women | Younger non-KP men | Older non-KP men | Female sex workers | Clients of female sex workers | Men who have sex with men |  |
| **Eastern and Southern Africa** |  |  |  |  |  |  |  |  |  |
| EMOD<sup>25</sup> | South Africa | Yes | Yes | Yes | Yes | Yes | Yes | NA | All |
| Goals<sup>26</sup> | South Africa | NA | NA | NA | NA | Yes | Yes | Yes | I<sub>1</sub>, I<sub>3</sub>, I<sub>4</sub> |
| Stone<sup>21</sup> | South Africa | NA<sup>a</sup> | NA | NA | NA | Yes | Yes | Yes | All |
| Thembisa<sup>28</sup> | South Africa | NA | NA | NA | NA | Yes | Yes | Yes | I<sub>1</sub>, I<sub>3</sub>, I<sub>4</sub> |
| Mishra<sup>29</sup> | South Africa, Eswatini and Lesotho combined | Yes | NA | Yes | NA | Yes | Yes | NA | All |
| Optima HIV<sup>27</sup> | South Africa, Eswatini, Mozambique, Malawi, and Zimbabwe | Yes | Yes | Yes | Yes | Yes | Yes | Yes | All |
| **Western and Central Africa** |  |  |  |  |  |  |  |  |  |
| Silhol Cameroon<sup>22</sup> | Yaoundé (Cameroon) | NA<sup>a</sup> | NA | NA | NA | Yes | Yes | Yes | All |
| Maheu-Giroux<sup>23</sup> | Côte d'Ivoire | Yes | Yes | Yes | Yes | Yes | Yes | Yes | I<sub>1</sub>, I<sub>3</sub>, I<sub>4</sub> |
| Silhol ATLAS<sup>30</sup> | Côte d'Ivoire, Mali, and Senegal | Yes | Yes | Yes | Yes | Yes | Yes | Yes | All |

<sup>a</sup>Stone and Silhol Cameroon: populations not included as the models are not age stratified.
NA: not available; Younger: aged 15-24 years; Older: aged 25+ years; KP: key populations (including female sex workers, their clients, and men who have sex with men);

### Analysis of the estimated indicators

We assessed the rank order of populations contributing to new infections identified by different indicators. For each indicator, we counted in how many models each population was ranked as the largest contributor and among the top 3 largest contributors for the year 2020 (and decade 2020-2029 for I_4_). This analysis was conducted on the 10 models that provided estimates for non-KP and FSW and was stratified by region (ESA and WCA) reflecting different epidemiological contexts.

Second, for each model and population (stratified by region), we evaluated the correlation between estimates of the contribution of each population from different indicators, for all 15 models, using correlation plots. We also calculated two ratios: i) between the *direct transmission* and *acquisition* indicators 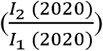, and ii) between 10-year *tPAF* and *direct transmission* indicators 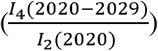, for each model and population, which can be interpreted as the magnitudes of differences i) in anticipated HIV prevention intervention impact when using a *direct transmission indicator* instead of an *acquisition indicator* and ii) when accounting for long-term secondary indirect transmissions instead of only capturing direct transmissions.

Third, we used Pearson’s correlation tests to assess the relationship between both indicator ratios 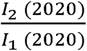 and 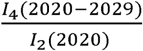 and key epidemiological factors (e.g., specific population size, HIV prevalence in 2020, intervention level, overall and by population). Finally, we evaluated to what extent the identified rank ordering of populations and the two indicator ratios changed from 2010 to 2020.

## Results

### Identification of priority populations across HIV indicators

When using the *acquisition indicator* (I_1_), five out of the six models in ESA identified non-KP women above age 25 years as the population among whom most infections occurred in 2020 due to their large size, but also identified younger non-KP women in the top 3 greatest contributors (with a higher incidence and a somewhat lower population size). Non-KP men above age 25 years were identified as acquiring the most infections only in the *Optima HIV* model for Zimbabwe (Figure 1a, Table S2a). In contrast, the *direct transmission indicator* (I_2_) more frequently (4/6 models) identified older non-KP men as the most important population (since women acquiring most infections implies that men directly transmit most infections) and identified older non-KP women as the population transmitting most infections in two models. Although CFSW in ESA were never among the three greatest contributors based on the *acquisition indicator* for 2020, they were the third largest contributor in two models based on all transmission indicators (I_2-4_; Figure 1b and Table S2a).

**Figure 1.**
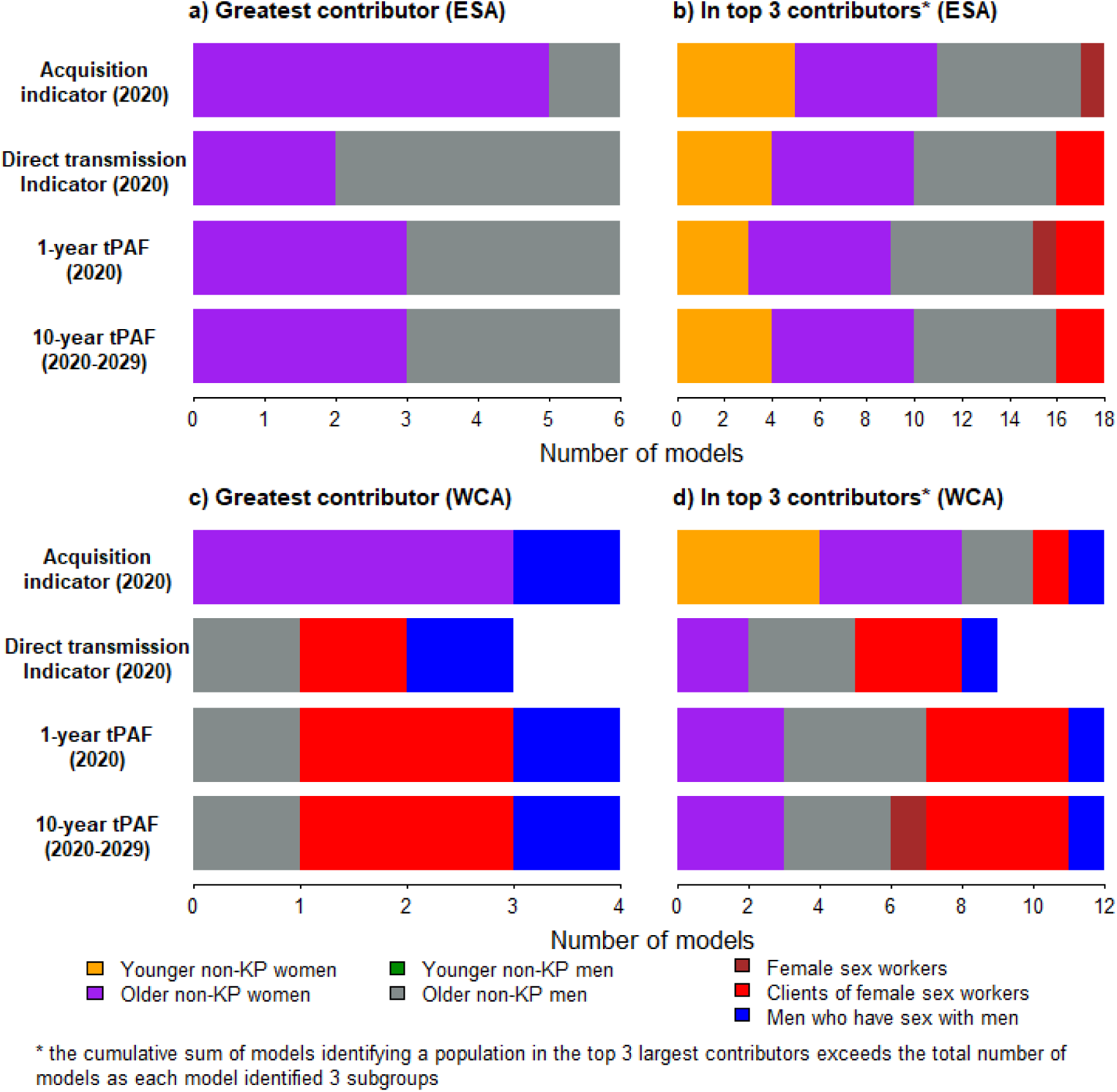
Differences in the populations identified as greatest (left panels) and among top 3 most important (right panels) contributors to new HIV infections by each indicator from mathematical models for a-b) Eastern and Southern Africa, and c-d) Western and Central Africa. I_1_: *acquisition indicator* (2020), I_2_: *direct transmission indicator* (2020), I_3_: 1-year transmission population-attributable fraction (2020), and I_4_: 10-year transmission population-attributable fraction (2020-2029). One model providing estimates for all selected populations in Western and Central Africa did not report estimates of the direct transmission indicator. None of the 10 models identified non-key population men aged 15-24 years old among the top 3 greatest contributors in any of the 4 indicators, whereas no model for Eastern and Southern Africa identified men who have sex with men among the top 3 greatest contributors in any of the 4 indicators. Additional details are provided in Table S2 (appendix pp 10-11). Younger: aged 15-24 years old; Older: aged 25+ years old; KP: key populations (including female sex workers, their clients, and men who have sex with men).

In WCA, despite the *acquisition indicator* (*I_1_*) identifying older non-KP women populations (3/4 models) as the greatest contributors to new infections, the three transmission indicators often identified CFSW, older non-KP men and MSM as the most important populations (Figure 1c, Table S2b). The 2020 *acquisition indicator* identified younger and older non-KP women among the three greatest contributors in all four models (Figure 1d, Table S2b).

### Differences between acquisition and transmission indicators

Almost all models estimated that younger and older non-KP women acquired substantially more infections than they transmitted in 2020. The *acquisition indicators* (I_1_) for these two populations were (median and min-max across model estimates) 1·4 (0·8-2·8) and 1·3 (0·9-1·7) times higher than *direct transmission indicator* (I_2_) in ESA (Figures 2a, S1a) and 3·3 (2·2-4·5) and 1·5 (1·2-2·6) times higher in WCA (Figures 3a, S1a), respectively. Although differences between *direct transmission* and *acquisition indicators* estimates for younger non-KP men in 2020 were mixed in ESA, they acquired 2·8-times (2·3-4·0) more infections than they transmitted in WCA (Figures 3a, S1a). Conversely, the 2020 *direct transmission indicator* was almost always larger than the 2020 *acquisition indicator* across models for older non-KP men 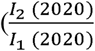 ratio of 1·4;1·0-1·8), CFSW (1·6; 0·9-3·6), and MSM (1·5; 0·7-3·1) in both regions, suggesting that these populations transmitted more infections than they acquired. Thus, preventing transmissions from them is likely to reduce new infections more than preventing acquisition (Figures 2a, 3a, S1a). Differences between *direct transmission* and *acquisition indicator* estimates for FSW were mixed for ESA 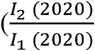 ratio of 0·8; 0·3-3·1), but most models for WCA predicted that they directly transmitted more infections than they acquired in 2020 (ratio of 1·1 (0·9-1·4), Figure S1a). The 10-year *tPAF* estimates for FSW (all regions combined) were 2·1-fold (0·8-5·3) higher than corresponding *acquisition indicator* estimates (Figures 2c,3c). Overall, relative differences between acquisition and all transmission indicators for non-KP were consistently larger in WCA compared to ESA, but similar for KPs in both regions (Figure S1a).

**Figure 2.**
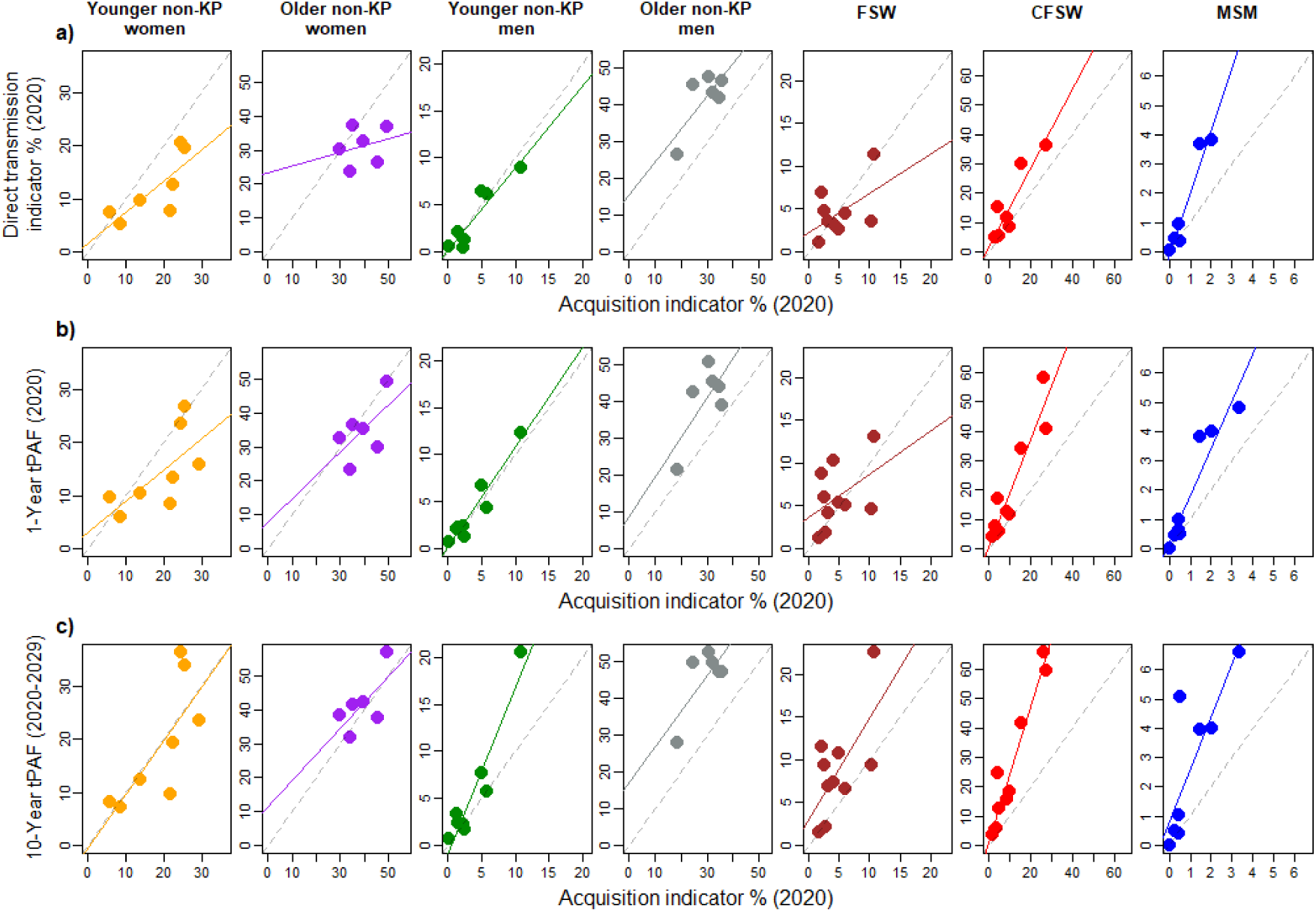
Correlation plots for Eastern and Southern Africa showing pairwise comparison of model estimates of a) *direct transmission indicator* (2020), b) 1-year *tPAF* (2020), and c) 10-year *tPAF* (2020-2029) on the y-axes with the *acquisition indicator* (2020) on x-axis, for each of the seven selected populations. Each point represents a pair of indicator estimates for a specific model/setting. Grey dashed diagonal lines indicate perfect agreement between indicator estimates. Solid coloured lines are regression lines from a linear model. Younger: aged 15-24 years old; Older: aged 25+ years old; FSW: female sex workers; CFSW: clients of female sex workers; MSM: men who have sex with men; KP: key populations (including female sex workers, their clients, and men who have sex with men).

**Figure 3.**
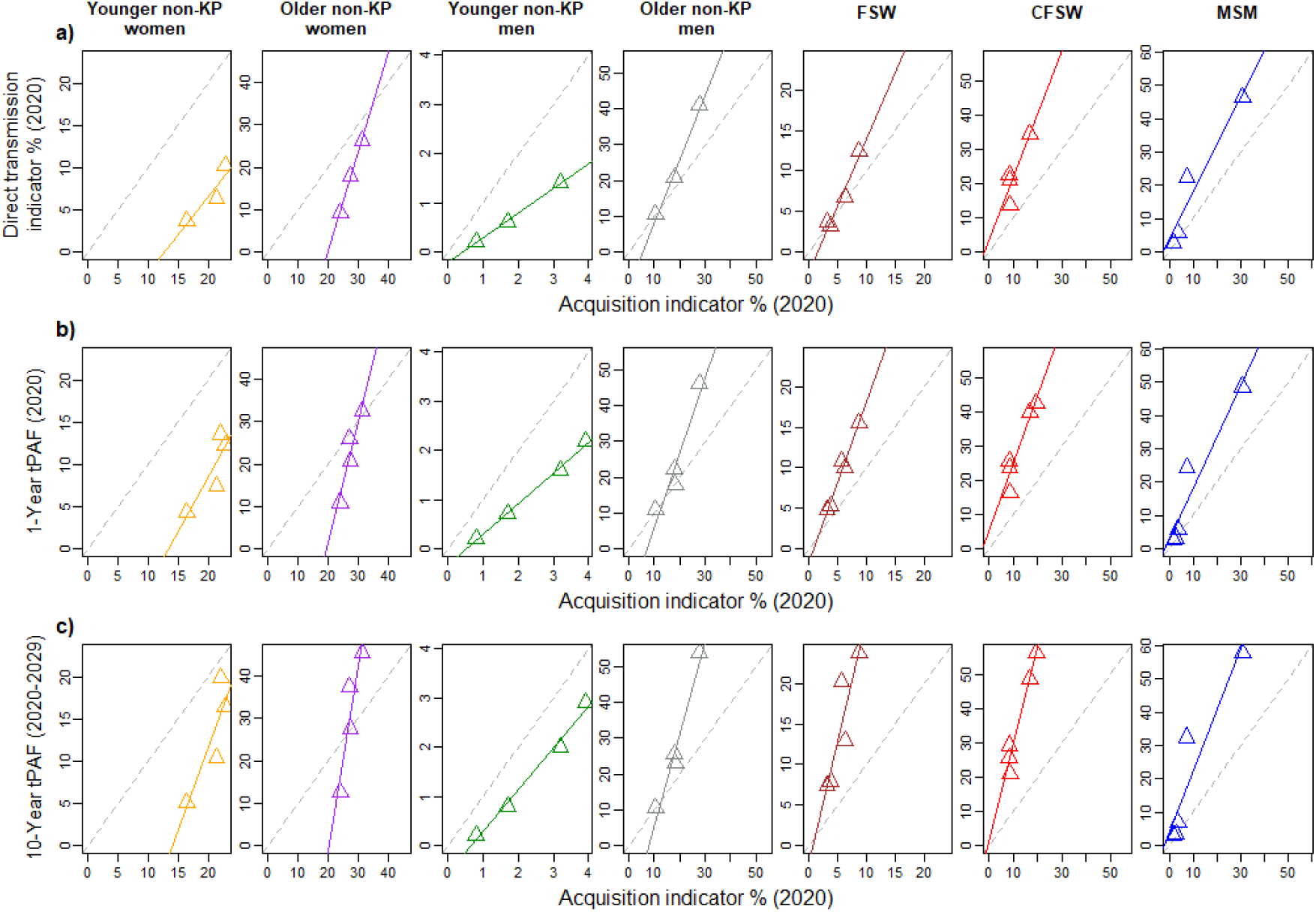
Correlation plots for Western and Central Africa showing pairwise comparison of model estimates of a) *direct transmission indicator* (2020), b) 1-year tPAF (2020), and c) 10-year tPAF (2020-2029) on the y-axes with the *acquisition indicator* (2020) on x-axis, for each of the seven selected populations. Each triangle represents a pair of indicator estimates for a specific model/setting. Grey dashed diagonal lines indicate perfect agreement between indicator estimates. Solid coloured lines are regression lines from a linear model. Younger: aged 15-24 years old; Older: aged 25+ years old; FSW: female sex workers; CFSW: clients of female sex workers; MSM: men who have sex with men; KP: key populations (including female sex workers, their clients, and men who have sex with men)

Magnitudes of differences were analogous when comparing 1-year *tPAF* and *acquisition indicators* across models and populations in 2020 as for comparing the *direct transmission* and the *acquisition indicators* (Figures 2b, 3b).

### Differences between transmission indicators

The 1-year tPAFs and *direct transmission indicators* across models were highly similar for all regions and populations (Figure 4a,b), but not always equal, particularly for FSW (the 1-year *tPAF* was 1·3-fold (1·1-3·1) larger). Furthermore, 10-year *tPAFs* for FSW (which accounts for more long-term secondary transmissions from clients and partners of populations) were 1·5-fold (0·7-2·0) larger than 1-year tPAFs, emphasising that large numbers of secondary infections could be averted in the long-term by preventing transmission from this population (Figures 4c, 4d). Finally, the differences between the 10-year *tPAFs* and the *direct transmissions indicator* was more substantial for younger women (median ratio of 1·5; 1·1-1·8) and especially FSW (2·0; 1·4-4·1) across models (Figure S1b).

**Figure 4.**
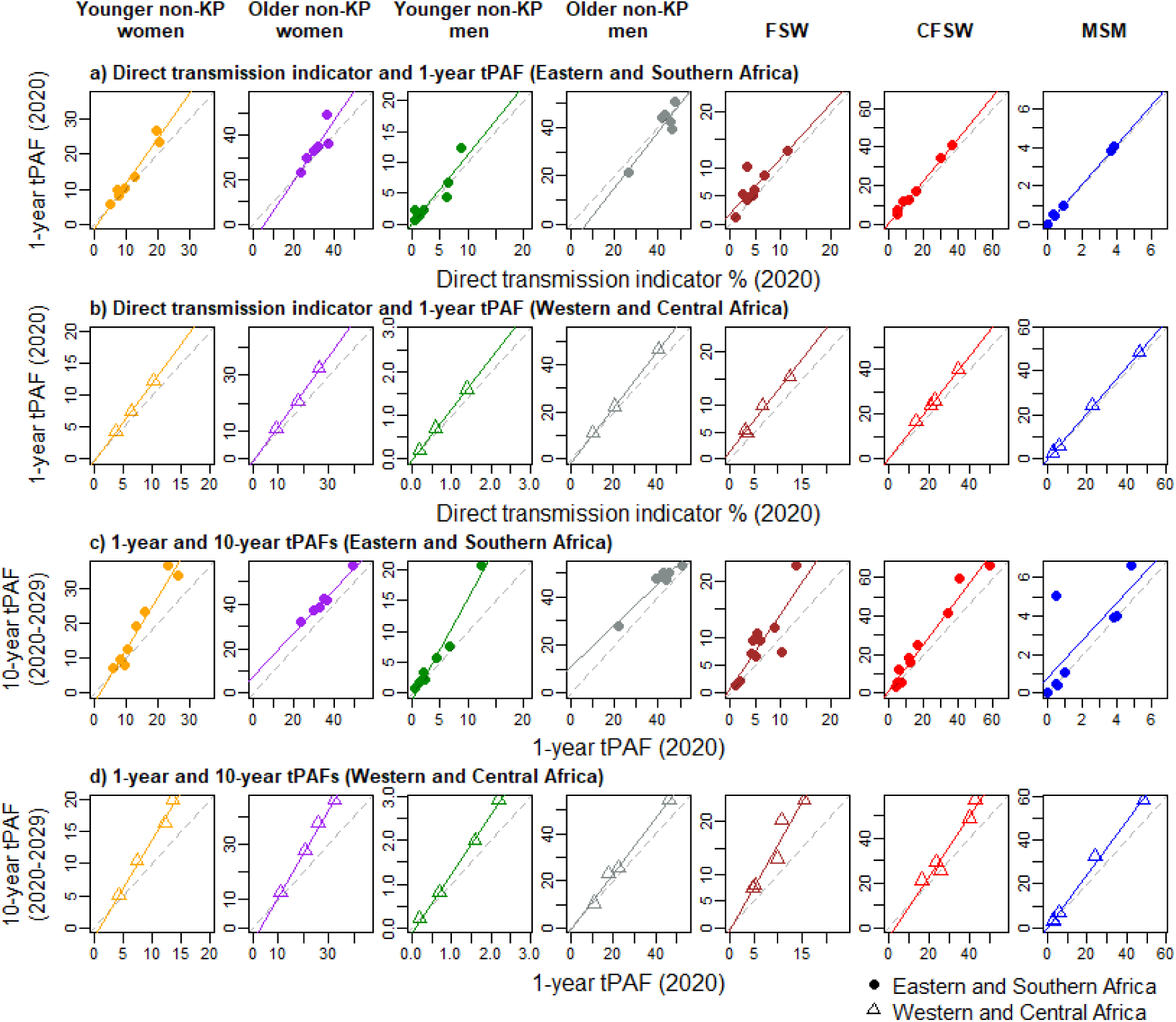
Correlation plots showing pairwise comparison of a-b) model estimates of 1-year *tPAF* with *direct transmission indicator* for 2020 as well as c-d) 10-year tPAF (2020-2029) with 1-year tPAF (2020), for each of the seven selected populations. Grey dashed diagonal lines indicate perfect agreement between indicator estimates. Solid coloured lines are regression lines from a linear model. Younger: aged 15-24 years old; Older: aged 25+ years old; FSW: female sex workers; CFSW: clients of female sex workers; MSM: men who have sex with men; KP: key populations (including female sex workers, their clients, and men who have sex with men)

### Factors influencing relative differences between indicators

A large estimated correlation coefficient suggests that younger non-KP women may transmit less infections than they acquired when overall HIV prevalence is low (see appendix Figures S2-S7 pp 13-23). Finally, the groups identified as priority populations and the 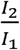 and indicator ratios were qualitatively similar when calculated for 2010 and 2020, suggesting 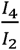 that differences in acquisition and transmission risks among populations have remained stable over this decade in these models (see appendix pp 24-26).

## Discussion

Our study showed that the choice of indicators can substantially influence which populations are identified as contributing most new HIV infections due to their unmet prevention and treatment needs. By ignoring transmissions, the *acquisition indicator* – by far the most commonly used indicator – often largely underestimates the contribution of men, and male key populations in particular, to new HIV infections. This could result in programs overlooking the important impact that prevention efforts in men can achieve^31,32^.

Additionally, our tPAF estimates identified that increasing treatment coverage among young women and female sex workers would not only avert direct transmissions to their partners and clients, but also similar numbers of secondary transmissions in the long term, an effect which was not captured by the direct transmission indicator.

In our model comparison, the *acquisition* and *transmission indicators* rarely identified the same priority populations or ranked them in the same order. For example, women were often identified as most important population by the *acquisition indicator*, compared to men when using the *direct transmission indicator,* which would dictate the type of intervention needed by each^33^. In WCA, older non-KP men, MSM, and especially CFSW – which have historically been neglected by HIV prevention programs in the region^34^ – were often identified as priority populations when using *transmission indicators* despite rarely being identified as such by the *acquisition indicator*. A possible reason for this result is the lower prevalence of HIV in WCA compared to ESA, which is associated with elevated concentration of the HIV acquisition risk among key and vulnerable populations compared to overall. This contrasted with estimates from models for ESA where non-KP were the largest contributors to acquisitions and transmissions due to much larger (∼6-fold) overall HIV prevalence among adults and less heterogeneity in HIV acquisition risks across populations compared to WCA^33^.

Although differences between *acquisition* and *direct transmission* indicators were expected, the magnitude of the difference was particularly large for young women (who acquire ∼1·6-fold more infections than they transmit), with their high incidence being likely due to increased biological susceptibility to HIV acquisition^19^, higher prevalence of STI elevating HIV acquisition risks to a larger extent than transmission risks^35^, the older age and lower HIV diagnosis and treatment coverage among their male partners^33,36^. These differences were again larger in WCA than ESA, partly due to the much higher coverage of male circumcision in WCA (reducing the acquisition risk among men but not a potential subsequent transmission to a female partner^37–39^). This supports recent efforts for reaching and diagnosing heterosexual men living with HIV through community safe spaces to prevent acquisitions among young women^40^. Larger estimates of *acquisition* than *direct transmission* indicators among younger non-KP were also influenced by ageing of these populations to the older age group, which does not provide them much time after acquiring the infection to subsequently transmit the infection. This effect was especially strong in WCA, where the relatively low rates of infection may cause new acquisitions to occur at an older age compared to ESA^33,39^.

Younger women and FSW are often considered priority populations by programs due to their disproportionate vulnerabilities and high HIV incidence. Although they were never identified as the population contributing the most by the *acquisition indicator* (or any other indicator), this was because of their smaller size (size of younger non-KP women and FSW: 1·5-4-fold and 20-190-fold smaller than older non-KP women across models, respectively), and younger women were nonetheless almost always among the top 3 population acquiring the most HIV infections.

Despite generally good agreement between the three selected *transmission indicators* when identifying priority populations, estimates of the magnitude of the contribution varied across indicators. Estimates of the contribution using 10-year *tPAFs*, which account for indirect secondary transmissions, instead of the *direct transmission indicator* were substantially (2-fold) higher for FSW. This highlights the importance of capturing long-term secondary onward transmissions from clients and partners of FSW to their other female partners to adequately evaluate the persisting importance of sex work in the current epidemic dynamics.

Our analysis has limitations. First, several models did not provide estimates for all of the seven selected populations and four indicators (Table 1), often because these groups were not modelled. The reason for models not representing MSM or non-KP groups was often because their contribution was assumed to be modest or their absence of age-stratification, respectively, which means that it did not meaningfully alter the model indicator estimates among KPs and by gender. As several modelling groups (e.g., *Goals* and *Thembisa* models) did not provide estimates for all populations represented in their model, we were only able to include a subset of all datasets (10/15 models) in our analysis identifying priority populations. Although modelling groups were requested to provide indicator estimates for transgender women, none of the models had included this population, due to large setting-specific data gaps^41^, however the contribution of this population to all new infections across indicators is likely to be limited by its modest size (around 0.1% of all adult women in Africa^13^). In practice, the calculation of the four indicators examined in our analysis should be complemented by assessments of the population vulnerability (e.g., “per capita” rate of HIV acquisition), its size, and the anticipated acceptability and adequacy of interventions when identifying “priority populations”, to not solely focus on contributions to all new infections. Finally, models represented sexual HIV acquisitions and transmissions among people aged 15 years and older, which means that transmissions through drug injection and vertical transmission were not included in our indicator calculations; including them could have increased estimates of the transmission indicators, especially among younger women.

Our study has several strengths. To our knowledge, this is the first model comparison looking at the identification of priority populations using different indicators, since almost all previous modelling analyses and population surveys measuring HIV impacts have solely focused on new acquisitions or prevalent infections^42,43^. Using 15 different mathematical model applications across Africa, with different designs and relying on different sets of assumptions, increased the robustness of our analysis and most of our findings, such as the disproportionate contribution of male populations to new transmissions, were consistent across most, if not all, settings. All models used for this analysis relied on reviews of country-specific data and reflected trends in country-specific HIV prevalence and intervention data, often by population and age. This allowed reflecting heterogeneities in HIV acquisition and transmission risks across populations. These heterogeneities in epidemic contexts partly explained differences between indicators, for example the larger differences between indicators for non-KP in WCA (for which very few mathematical models exist) compared to ESA, potentially due to higher coverage of male circumcision. Finally, having indicator estimates for 2010 and 2020 enabled us to show that although indicator levels might have changed over time for specific populations, indicator differences appeared to have remained stable over the recent period.

Transmission indicators are often complex to calculate because they require both local data and assumptions, and careful model calibration. Nevertheless, they provide an opportunity to characterise HIV incidence and improve upon how decisions and interventions can reduce HIV transmission, and hence complement the widely used and cited *acquisition indicator* measuring the risk of HIV due to remaining prevention gaps. As a result, epidemiologic assessments using mathematical models should systematically include both transmission indicators and acquisition indicators, with the 10-year transmission population-attributable fractions (*tPAFs*) better reflecting the potential long-term impacts of interventions reducing HIV transmission. This information better reflects the complexity of HIV, and can be used to decide which interventions would more meaningfully impact new infections (i.e. interventions decreasing HIV acquisition risk, such as increasing PrEP access, or reducing transmission, such as increasing testing). Here, epidemic models applied across diverse African HIV epidemic settings provide consistent evidence of the potential disproportionate epidemiologic impacts of men and key populations reinforcing the need for well specified interventions with the goal of optimising the impact of HIV prevention and treatment programs^44^.

## Data Availability

The model estimates that support the findings of this analysis are available from the corresponding author upon reasonable request, after consultation with the modelling teams.

## Contributors

JSta, OS, DDi, SB, JWI-E, and M-CB conceptualised the study. JSta, JWI-E and M-CB designed the spreadsheet used to collate the different model estimates and wrote its supporting documentation (including the indicator definitions and a survey measuring indicators use and interpretations), with input from RSi, KMM, and DDi. Data collation was coordinated by RSi, RDB, and JSta, with guidance from KMM, JWI-E and M-CB. The models used for this analysis were developed, calibrated and/or analysed by AB, H-YK (EMOD), JStov (Goals), JSton, PV (Stone), LFJ (Thembisa), SM (Mishra), SLK, RM-H, RSt, DPW (Optima HIV), RS (Silhol Cameroon), MM-G (Maheu-Giroux), RS, MM-G (Silhol ATLAS). The analysis and statistics were computed by RSi and RDB, with input from M-CB, with the other authors providing further contributions to the analysis description and interpretation of the results.

RSi and RDB wrote the first version of the manuscript, which was revised by KMM, DDi, JWI-E, and M-CB. All authors had full access to all the data in the analysis, reviewed manuscript drafts, read and approved the final version of the manuscript, and had final responsibility for the decision to submit for publication.

## Acknowledgements

Supported partly by UNAIDS and the HPTN Modelling Centre, which is funded by the U.S. National Institutes of Health (NIH UM1 AI068617) through HPTN. RSi, RDB, KMM, JSta, OS, JWI-E and M-CB acknowledge funding from the MRC Centre for Global Infectious Disease Analysis (reference MR/X020258/1), funded by the UK Medical Research Council (MRC). This UK funded award is carried out in the frame of the Global health EDCTP3 Joint Undertaking. RSi, JSton, PV, M-CB and MM-G acknowledge funding from the Wellcome Trust (WT 226619/Z/22/Z). MM-G and SM’s research program are supported by Tier 2 *Canada Research Chairs* in *Population Health Modeling* and *Mathematical Modeling and Program Science,* respectively. AB was supported by the US National Institutes of Health (NIH R01AI179417). Support for EMOD-HIV and Thembisa was provided by the Gates Foundation. Country models utilized for Optima HIV have been supported by the Gates Foundation, Global Fund, and the World Bank. For the purpose of open access, the author has applied a Creative Commons Attribution (CC BY) license to any Author Accepted Manuscript version arising.

The authors would like to thank UNAIDS and the collaborators involved in the mathematical modelling, as well as the participants of the empirical studies on which the models rely.

