## Supplement for "Identifying priority populations for HIV interventions using acquisition and transmission indicators: a combined analysis of 15 mathematical models from 10 African countries"

### Methods: models used for the analysis

Table S1. Characteristics of the models of HIV transmission and modelled epidemic contexts used for this analysis (median estimates across model simulations).

| Model Name (Model Type) | Countries (region) | Populations included | Calibration data | Population | Relative size of population in 2020 | HIV prevalence in 2020 | Viral suppression among adults with HIV in 2020 |
|---|---|---|---|---|---|---|---|
| *EMOD* (stochastic individual-based) [1] | South Africa (ESA) | YW, OW, YM, OM, FSW CFSW | Prevalence by age/sex, incidence by age/sex, population by age/sex, number on ART by sex, ART coverage by age/sex. | All | 100% | 18.2% | 54.6% |
| | | | | YW | 12.7% | 9.1% | 33.7% |
| | | | | OW | 39.1% | 26.1% | 58.0% |
| | | | | YM | 12.0% | 2.7% | 29.6% |
| | | | | OM | 33.4% | 16.3% | 55.3% |
| | | | | FSW | 0.2% | 55.3% | 27.2% |
| | | | | CFSW | 2.6% | 38.7% | 48.5% |
| *Goals* (deterministic compartmental) [2] | South Africa (ESA) | YW, OW, YM, OM, FSW CFSW, MSM | HIV prevalence All, FSW and MSM<br>Historical ART numbers | All | 100% | 26.6% | 56.0%[a] |
| | | | | FSW | 0.4% | 43.7% | 33.0%[a] |
| | | | | CFSW | 1.8% | 32.6% | 68.0%[a] |
| | | | | MSM | 0.9% | 22.2% | 68.0%[a] |
| *Stone* (deterministic compartmental) [3] | South Africa (ESA) | FSW CFSW, MSM (non-MSM population not age-structured) | Relative sizes of key populations<br>HIV prevalence and ART coverage among adults by sex, FSW, CFSW and MSM | All | 100% | 19.0% | 62.9% |
| | | | | FSW | 0.4% | 55.1% | 53.5% |
| | | | | CFSW | 13.3% | 20.8% | 46.6% |
| | | | | MSM | 1.0% | 37.6% | 25.8% |
| *Thembisa* (deterministic compartmental) [4] | South Africa (ESA) | YW, OW, YM, OM, FSW | HIV prevalence in MSM and FSW; HIV prevalence in | All | 100% | 15.8% | 62.9% |
| | | | | YW | 11.2% | 9.4% | 48.9% |
| | | | | OW | 40.6% | 24.8% | 66.6% |

| | | | | | | | |
|---|---|---|---|---|---|---|---|
| | | CFSW, MSM | antenatal clinic surveys (by age) HIV prevalence in household surveys (by age and sex) Recorded death data (by age and sex) ART: males, females, children HIV testing history: males, females, children, stratified by age | YM | 8.3% | 2.5% | 63.9% |
| | | | | OM | 25.1% | 5.1% | 55.9% |
| | | | | FSW | 0.3% | 50.5% | 56.5% |
| | | | | CFSW | 13.8% | 33.6% | 59.8% |
| | | | | MSM | 0.7% | 24.1% | 46.4% |
| *Mishra* (deterministic compartmental) [5] | South Africa, Lesotho and Eswatini combined (ESA) | YW, OW, YM, OM, FSW CFSW | HIV prevalence by sex/age-group combinations, among FSW by age-group. HIV prevalence ratios FSW: non-FSW females, by age-group HIV prevalence ratios clients: non-client males, age-standardized ART coverage over time overall & by risk and age-group combinations | All | 100% | 19.2% | 60.5% |
| | | | | YW | 9.3% | 11.3% | 54.1% |
| | | | | OW | 38.6% | 3.9% | 74.3% |
| | | | | YM | 8.6% | 3.2% | 34.5% |
| | | | | OM | 26.4% | 12.1% | 47.4% |
| | | | | FSW | 0.9% | 46.9% | 20.3% |
| | | | | CFSW | 16.2% | 17.2% | 47.1% |
| *Optima HIV* (deterministic compartmental) [6] | South Africa (ESA) | YW, OW, YM, OM, FSW CFSW, MSM | Population size, HIV prevalence by population, treatment, PLHIV, new HIV infections, HIV-related deaths, HIV diagnoses. HIV testing (diagnosis), ART, viral load monitoring (viral suppression) | All | 100% | 17.0% | 65.9% |
| | | | | YW | 11.2% | 16.5% | 67.1% |
| | | | | OW | 38.3% | 20.1% | 66.2% |
| | | | | YM | 11.3% | 7.5% | 67.6% |
| | | | | OM | 35.7% | 15.9% | 65.1% |
| | | | | FSW | 0.3% | 37.6% | 65.0% |
| | | | | CFSW | 2.7% | 14.0% | 66.0% |
| | | | | MSM | 0.7% | 30.4% | 56.9% |
| | | | | All | 100% | 25.0% | 69.9% |

| | | | | | | | |
|---|---|---|---|---|---|---|---|
| *Optima HIV* (deterministic compartmental) [6] | Eswatini (ESA) | YW, OW, YM, OM, FSW CFSW, MSM | Population size, HIV prevalence by population, treatment, PLHIV, new HIV infections, HIV-related deaths, HIV diagnoses. HIV testing (diagnosis), ART, viral load monitoring (viral suppression) | YW | 16.2% | 6.9% | 65.9% |
| | | | | OW | 33.4% | 39.4% | 70.6% |
| | | | | YM | 15.8% | 4.8% | 68.0% |
| | | | | OM | 31.8% | 28.8% | 69.9% |
| | | | | FSW | 0.6% | 53.3% | 62.3% |
| | | | | CFSW | 1.8% | 27.2% | 68.1% |
| | | | | MSM | 0.4% | 11.5% | 74.6% |
| *Optima HIV* (deterministic compartmental) [6] | Zimbabwe (ESA) | YW, OW, YM, OM, FSW CFSW, MSM | Population size, HIV prevalence by population, treatment, PLHIV, new HIV infections, HIV-related deaths, HIV diagnoses. HIV testing (diagnosis), ART, viral load monitoring (viral suppression) | All | 100% | 14.3% | 70.7% |
| | | | | YW | 18.2% | 6.1% | 63.3% |
| | | | | OW | 36.1% | 20.1% | 72.8% |
| | | | | YM | 15.1% | 2.9% | 69.3% |
| | | | | OM | 25.9% | 17.6% | 69.9% |
| | | | | FSW | 0.5% | 41.2% | 66.2% |
| | | | | CFSW | 3.9% | 17.0% | 68.1% |
| | | | | MSM | 0.3% | 20.4% | 71.8% |
| *Optima HIV* (deterministic compartmental) [6] | Mozambique (ESA) | YW, OW, YM, OM, FSW CFSW, MSM | Population size, HIV prevalence by population, treatment, PLHIV, new HIV infections, HIV-related deaths, HIV diagnoses. HIV testing (diagnosis), ART, viral load monitoring (viral suppression) | All | 100% | 7.7% | 18.7% |
| | | | | YW | 17.1% | 5.8% | 14.9% |
| | | | | OW | 32.1% | 11.0% | 19.4% |
| | | | | YM | 16.1% | 3.7% | 16.2% |
| | | | | OM | 28.4% | 7.2% | 20.0% |
| | | | | FSW | 1.5% | 11.1% | 16.6% |
| | | | | CFSW | 4.1% | 6.9% | 19.6% |
| | | | | MSM | 0.6% | 7.2% | 25.7% |
| *Optima HIV* (deterministic compartmental) [6] | Malawi (ESA) | YW, OW, YM, OM, FSW CFSW, MSM | Population size, HIV prevalence by population, treatment, PLHIV, new HIV infections, HIV-related deaths, HIV diagnoses. | All | 100% | 8.8% | 74.7% |
| | | | | YW | 19.1% | 2.6% | 75.1% |
| | | | | OW | 33.6% | 14.3% | 77.3% |
| | | | | YM | 17.1% | 1.6% | 81.7% |
| | | | | OM | 26.8% | 10.1% | 70.7% |
| | | | | FSW | 0.3% | 49.6% | 63.3% |

| | | | | | | | |
|---|---|---|---|---|---|---|---|
| | | | HIV testing (diagnosis), ART, viral load monitoring (viral suppression) | CFSW<br>MSM | 2.6%<br>0.4% | 10.1%<br>12.8% | 68.8%<br>73.9% |
| *Silhol Yaoundé* (deterministic compartmental) [7] | Yaoundé (Cameroon, WCA) | FSW CFSW, MSM (non-MSM population not age-structured) | Size of key populations. HIV prevalence among all females, all males, and key populations. ART coverage by gender, viral suppression coverage in each group | All<br>FSW<br>CFSW<br>MSM | 100%<br>0.7%<br>8.0%<br>0.9% | 3.9%<br>13.9%<br>3.5%<br>31.1% | 42.7%<br>69.9%<br>37.4%<br>39.6% |
| *Maheu-Giroux* (deterministic compartmental) [8] | Côte d'Ivoire (WCA) | YW, OW, YM, OM, FSW CFSW, MSM | Size of key populations, HIV prevalence by risk group and age, ART coverage | All<br>YW<br>OW<br>YM<br>OM<br>FSW<br>CFSW<br>MSM | 100%<br>19.4%<br>28.3%<br>17.4%<br>25.8%<br>0.8%<br>7.6%<br>0.6% | 2.3%<br>1.2%<br>3.5%<br>0.2%<br>1.8%<br>10.5%<br>5.4%<br>12.4% | 42.6%<br>35.1%<br>46.6%<br>24.9%<br>41.7%<br>40.8%<br>39.0%<br>48.5% |
| *Silhol ATLAS* (deterministic compartmental) [9] | Côte d'Ivoire (WCA) | YW, OW, YM, OM, FSW CFSW, MSM | Size of key populations. HIV prevalence among all females, all males, and key populations. HIV incidence among all females and all males/ Number of AIDS deaths. ART and viral suppression coverage. Fraction of PLHIV diagnosed in each group. Number of HIV tests each year, and fraction of positive tests. | All<br>YW<br>OW<br>YM<br>OM<br>FSW<br>CFSW<br>MSM | 100%<br>19.2%<br>27.4%<br>18.2%<br>26.4%<br>0.7%<br>7.3%<br>0.7% | 2.4%<br>1.2%<br>4.0%<br>0.2%<br>2.4%<br>9.1%<br>2.7%<br>8.7% | 51.8%<br>43.2%<br>60.4%<br>20.9%<br>46.7%<br>49.6%<br>45.3%<br>41.5% |

| | | | | | | | |
|---|---|---|---|---|---|---|---|
| *Silhol ATLAS* (deterministic compartmental) [9] | Mali (WCA) | YW, OW, YM, OM, FSW CFSW, MSM | Size of key populations. HIV prevalence among all females, all males, and key populations. HIV incidence among all females and all males/ Number of AIDS deaths. ART and viral suppression coverage. Fraction of PLHIV diagnosed in each group. Number of HIV tests each year, and fraction of positive tests. | All<br>YW<br>OW<br>YM<br>OM<br>FSW<br>CFSW<br>MSM | 100%<br>20.5%<br>30.3%<br>17.4%<br>26.1%<br>0.3%<br>5.3%<br>0.2% | 0.6%<br>0.3%<br>0.9%<br><0.1%<br>0.4%<br>9.1%<br>2.0%<br>14.3% | 33.3%<br>23.0%<br>39.1%<br>8.1%<br>25.6%<br>36.6%<br>28.3%<br>42.7% |
| *Silhol ATLAS* (deterministic compartmental) [9] | Senegal (WCA) | YW, OW, YM, OM, FSW CFSW, MSM | Size of key populations. HIV prevalence among all females, all males, and key populations. HIV incidence among all females and all males/ Number of AIDS deaths. ART and viral suppression coverage. Fraction of PLHIV diagnosed in each group. Number of HIV tests each year, and fraction of positive tests. | All<br>YW<br>OW<br>YM<br>OM<br>FSW<br>CFSW<br>MSM | 100%<br>21.2%<br>18.9%<br>30.2%<br>26.9%<br>0.3%<br>2.0%<br>0.2% | 0.3%<br>0.1%<br>0.5%<br><0.1%<br>0.2%<br>4.2%<br>1.8%<br>21.6% | 61.0%<br>58.6%<br>74.7%<br>40.8%<br>59.9%<br>43.8%<br>60.4%<br>24.5% |

[a] ART coverage among adults living with HIV in 2020

ESA: Eastern and Southern Africa; WCA: Western and Central Africa

YW: non-KP women aged 15-24 years old; OW: non-KP women aged 25-49 years old; YM: non-KP men aged 15-24 years old; OM: non-KP men aged 25-49 years old; FSW: female sex workers aged 15-49 years old; CFSW: clients of female sex workers aged 15-49 years old; MSM: men who have sex with men aged 15-49 years old.

KP: key populations (including FSW, CFSW, and MSM)

### Methods: definitions of the four HIV contribution indicators

A*cquisition indicator I₁* estimates the fraction of all newly acquired infections in one year that are among each specific population. It is calculated as the fraction ($Acq_Y^j$) of all new infections over the year Y ($Inf_Y$) which were acquired by members of the population $j$ from any of their partners:

$$Acq_Y^j = \frac{\sum_i T_{i \to j_Y}}{Inf_Y} \times 100$$

Where $T_{i \to j_Y}$ is the cumulative number of HIV infections transmitted from members of population *i* to members of population *j* in year Y.

*Direct transmission indicator $I_2$* is the fraction of newly transmitted infections in one year that are from each population. It is calculated as the fraction ($Tra_Y^i$) of all new infections in year Y ($Inf_Y$) which were directly transmitted by members of population $i$ to any of their partners:

$$Tra_Y^i = \frac{\sum_j T_{i \to j_Y}}{Inf_Y} \times 100$$

The *transmission population attributable fraction (tPAF) over 1 and 10 years ($I_3$ and $I_4$)* measure the fraction of all new infections over a time period *dt* (i.e., in year Y or over a decade as in our analysis) which could be averted by blocking all transmissions from a specific population over the period (but not their risk of acquiring HIV)[10,11]. It is calculated as the relative difference between the number of all new infections ($Inf_{dt}$) in a previously calibrated baseline scenario, allowing all transmissions over a period *dt*, and the number of new infections ($Inf_{dt}^i$) in a counterfactual scenario where the probability of transmissions from a population *i* is zero during *dt*. The difference $Inf_{dt} - Inf_{dt}^i$ is the number of new infections that can be directly or indirectly *attributed* to the population *i* over *dt*.

$$tPAF_{dt}^i = \left(\frac{Inf_{dt} - Inf_{dt}^i}{Inf_{dt}}\right) \times 100$$

Because blocking all transmissions from population *i* to population *j* in the counterfactual scenario may also avert future subsequent transmissions from population *j* to their partners, the transmission indicator *tPAF* reflects the long-term overall impact of hypothetically preventing transmission from a population *i* [11]. The sum of the *tPAF* across different populations can exceed 100% as individuals can acquire HIV from different populations.

### Supplement results: indicator estimates for 2020 and 2020-2029

Table S2. Populations identified as priority populations for additional HIV prevention and treatment efforts (i.e., in the top three greatest contributors to new HIV infections) according to our four selected indicators, for the 10 epidemic models providing estimates for each selected population. The three columns on the right report levels of agreement between indicators ("yes" (green): the identified the same three populations and in the same order; "partial" (amber): identified the same three populations but in different order; "no" (red): the three populations identified are different)

| | Rank | $I_1$: Acquisition indicator (2020) | $I_2$: Direct transmission indicator (2020) | $I_3$: 1-year tPAF (2020) | $I_4$: 10-year tPAF (2020-2029) | Agreements between indicators | | |
|---|---|---|---|---|---|---|---|---|
| | | | | | | $I_1$ and $I_2$ | $I_2$ and $I_3$ | $I_3$ and $I_4$ |
| **a) Eastern and Southern Africa** | | | | | | | | |
| *EMOD HIV* (South Africa) | 1st | OW | OM | OM | OM | Partial | Yes | Yes |
| | 2nd | OM | OW | OW | OW | | | |
| | 3rd | YW | YW | YW | YW | | | |
| *Optima* (South Africa) | 1st | OW | OM | OM | OM | Partial | Yes | Yes |
| | 2nd | OM | OW | OW | OW | | | |
| | 3rd | YW | YW | YW | YW | | | |
| *Optima* (Eswatini) | 1st | OW | OM | OW | OW | Partial | No | No |
| | 2nd | OM | OW | OM | OM | | | |
| | 3rd | YW | YW | FSW | YW | | | |
| *Optima* (Zimbabwe) | 1st | OM | OW | OW | OW | No | Yes | Yes |
| | 2nd | OW | OM | OM | OM | | | |
| | 3rd | YW | CFSW | CFSW | CFSW | | | |
| *Optima* (Mozambique) | 1st | OW | OW | OW | OW | Partial | Partial | Yes |
| | 2nd | YW | OM | YW | YW | | | |
| | 3rd | OM | YW | OM | OM | | | |
| *Optima* (Malawi) | 1st | OW | OM | OM | OM | Partial | Yes | Yes |

| | | | | | | | | |
|---|---|---|---|---|---|---|---|---|
| | 2nd | OM | OW | OW | OW | | | |
| | 3rd | FSW | CFSW | CFSW | CFSW | | | |
| **b) Western and Central Africa** | | | | | | | | |
| *Maheu-Giroux* (Côte d'Ivoire) | 1st | OW | . | CFSW | CFSW | NA | NA | Yes |
| | 2nd | YW | N.A. | OW | OW | | | |
| | 3rd | CFSW | | OM | OM | | | |
| *Silhol ATLAS* (Côte d'Ivoire) | 1st | OW | OM | OM | OM | No | Yes | Yes |
| | 2nd | OM | OW | OW | OW | | | |
| | 3rd | YW | CFSW | CFSW | CFSW | | | |
| *Silhol ATLAS* (Mali) | 1st | OW | CFSW | CFSW | CFSW | No | Yes | Partial |
| | 2nd | YW | OM | OM | OW | | | |
| | 3rd | OM | OW | OW | OM | | | |
| *Silhol ATLAS* (Senegal) | 1st | MSM | MSM | MSM | MSM | No | Yes | No |
| | 2nd | OW | CFSW | CFSW | CFSW | | | |
| | 3rd | YW | OM | OM | FSW | | | |

YW: non-KP women aged 15-24 years old; OW: non-KP women aged 25-49 years old; YM: non-KP men aged 15-24 years old; OM: non-KP men aged 25-49 years old; FSW: female sex workers aged 15-49 years old; CFSW: clients of female sex workers aged 15-49 years old; MSM: men who have sex with men aged 15-49 years old. KP: key populations (including FSW, CFSW, and MSM); NA: estimate not available

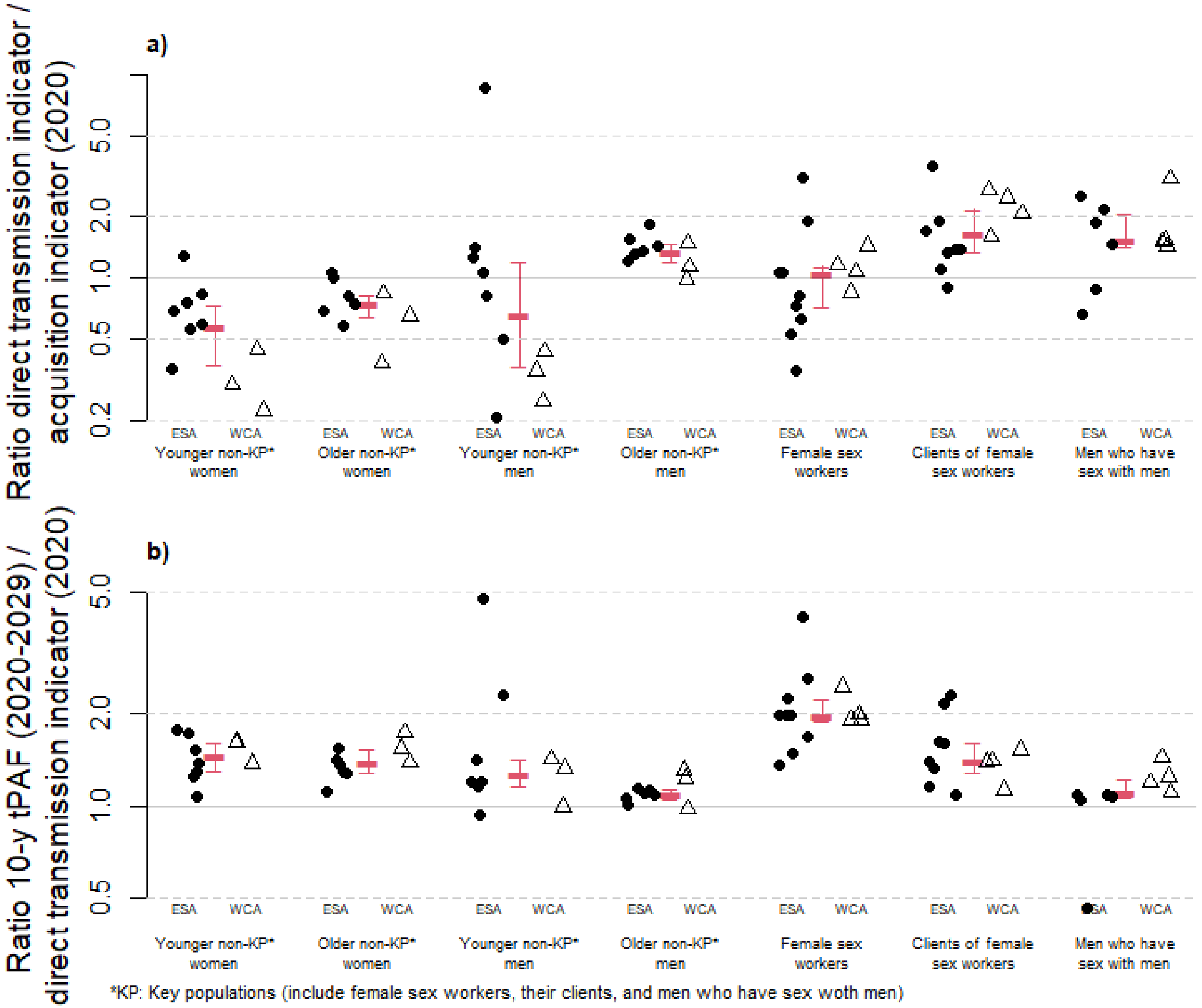

Figure S1. Ratios (shown in logscale) between model estimated a) *direct transmission indicator* over the *acquisition indicator* for 2020 ($\frac{I_2(2020)}{I_1(2020)}$) , and b) 10-year tPAF (2020-2029) over the *direct transmission indicator* for 2020 ($\frac{I_4(2020-2029)}{I_2(2020)}$), for the seven selected populations, stratified by African region (dots=Eastern and Southern Africa (ESA), triangles = Western and Central Africa (WCA)). Red bars and intervals correspond to the median and interquartile range of ratio estimates for the populations (all regions). Younger: aged 15-24 years old; Older: aged 25+ years old; KP: key populations (including female sex workers, their clients, and men who have sex with men)

### Sensitivity analysis: epidemiological factors influencing relative differences between indicators

A specific analysis aimed at evaluating under which overall or population-specific epidemiological context would our selected contribution indicators differ the most. To achieve this, we assessed the relationship between the indicator ratios $\frac{I_2\ (2020)}{I_1\ (2020)}$ and $\frac{I_4(2020-2029)}{I_2(2020)}$ and key epidemiological factors from all models: overall and population-specific HIV prevalence and incidence rates, incidence/prevalence ratios, fractions of all PLHIV who have a suppressed viral load, the coefficient of variations across populations for each of these previous four outcomes (which represents the degree of heterogeneity across HIV risk and treatment coverage across populations within the same setting), the relative size of the population (relative to all adults aged 15+ years old), ratio of the coverage of HIV viral suppression in the populations compared to overall (i.e., all adults aged 15+ years old), and fraction of PLHIV with an unsuppressed viral load who are members of the population (i.e., distribution of unsuppressed HIV). We also calculated the correlation coefficients of the indicator ratios for FSW with the model assumed average duration of sex work. Test p-values were adjusted using the Benjamini-Hochberg procedure to account for the high number of comparisons[12], and we reported correlations with the highest absolute correlations coefficients.

There was a non-significant positive correlation between overall HIV prevalence and the $\frac{I_2}{I_1}$ ratio for younger non-KP women, suggesting that young women may transmit much less infections than they acquired when HIV prevalence was low (Pearson's rho=0.78; adjusted p-value=0.4; Figure S3a). Larger fractions of younger non-KP men among all PLHIV with an unsuppressed HIV viral load was positively associated with larger $\frac{I_4}{I_2}$ ratios for this population, however this correlation was highly influenced by a single model estimate (rho=0.83; p=0.27; Figure S6o). There was a positive correlation between the coefficient of variation in viral suppression across populations and the $\frac{I_4}{I_2}$ ratio for younger non-KP women, suggesting larger number of secondary transmissions stemming from unmet treatment needs of young women (represented by large $\frac{I_4}{I_2}$ ratios) occurred when there were important heterogeneities in the levels of viral suppression across modelled populations (rho=0.76; p=0.27; Figure S6h).

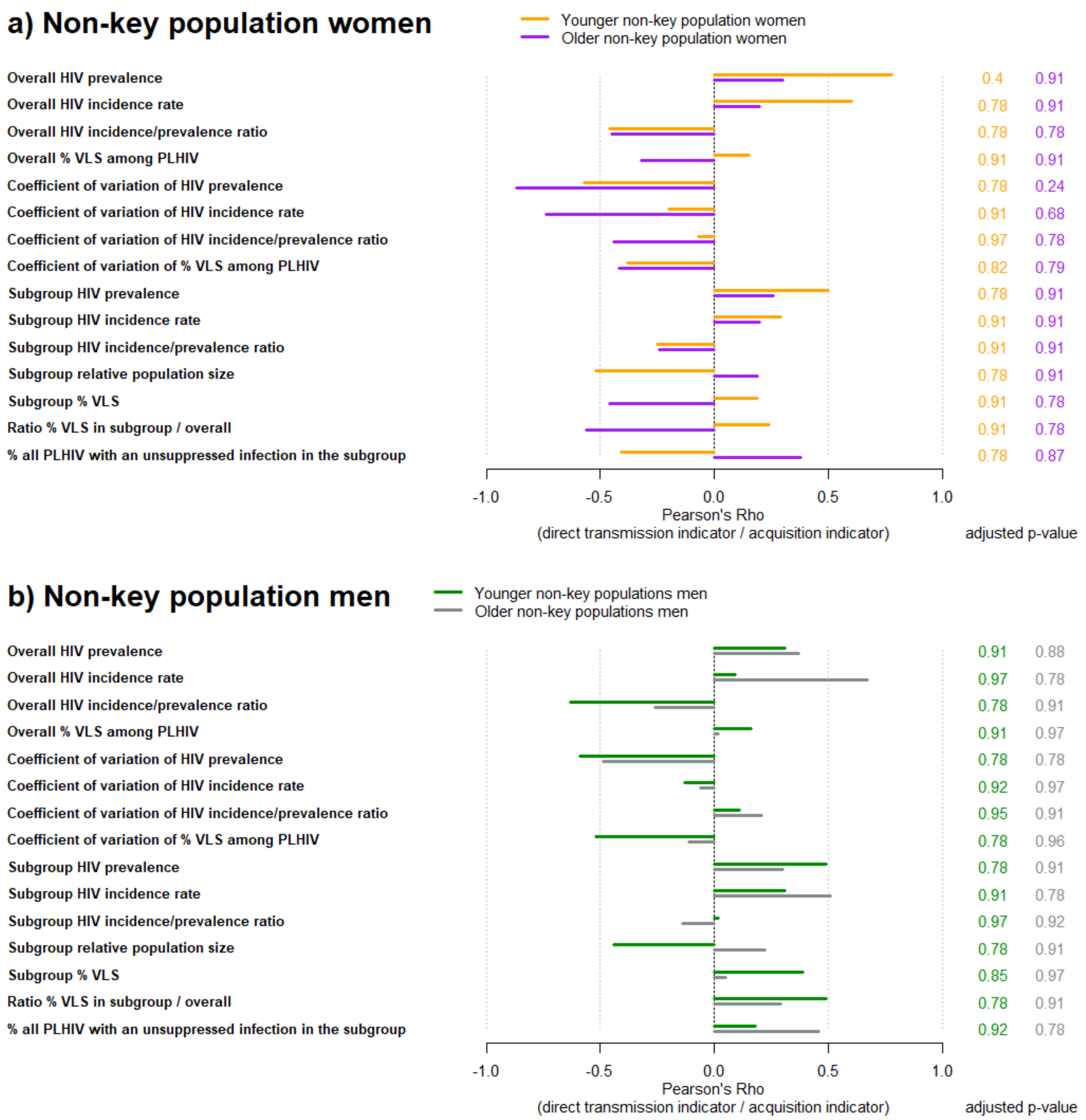
a) Non-key population women
Younger non-key population women
Older non-key population women
Overall HIV prevalence
Overall HIV incidence rate
Overall HIV incidence/prevalence ratio
Overall % VLS among PLHIV
Coefficient of variation of HIV prevalence
Coefficient of variation of HIV incidence rate
Coefficient of variation of HIV incidence/prevalence ratio
Coefficient of variation of % VLS among PLHIV
Subgroup HIV prevalence
Subgroup HIV incidence rate
Subgroup HIV incidence/prevalence ratio
Subgroup relative population size
Subgroup % VLS
Ratio % VLS in subgroup / overall
% all PLHIV with an unsuppressed infection in the subgroup
0.4 0.91
0.78 0.91
0.78 0.78
0.91 0.91
0.78 0.24
0.91 0.68
0.97 0.78
0.82 0.79
0.78 0.91
0.91 0.91
0.91 0.91
0.78 0.91
0.91 0.78
0.91 0.78
0.78 0.87
-1.0
-0.5
0.0
0.5
1.0
Pearson's Rho
(direct transmission indicator / acquisition indicator)
adjusted p-value
b) Non-key population men
Younger non-key populations men
Older non-key populations men
Overall HIV prevalence
Overall HIV incidence rate
Overall HIV incidence/prevalence ratio
Overall % VLS among PLHIV
Coefficient of variation of HIV prevalence
Coefficient of variation of HIV incidence rate
Coefficient of variation of HIV incidence/prevalence ratio
Coefficient of variation of % VLS among PLHIV
Subgroup HIV prevalence
Subgroup HIV incidence rate
Subgroup HIV incidence/prevalence ratio
Subgroup relative population size
Subgroup % VLS
Ratio % VLS in subgroup / overall
% all PLHIV with an unsuppressed infection in the subgroup
0.91 0.88
0.97 0.78
0.78 0.91
0.91 0.97
0.78 0.78
0.92 0.97
0.95 0.91
0.78 0.96
0.78 0.91
0.91 0.78
0.97 0.92
0.78 0.91
0.85 0.97
0.78 0.91
0.92 0.78
-1.0
-0.5
0.0
0.5
1.0
Pearson's Rho
(direct transmission indicator / acquisition indicator)
adjusted p-value

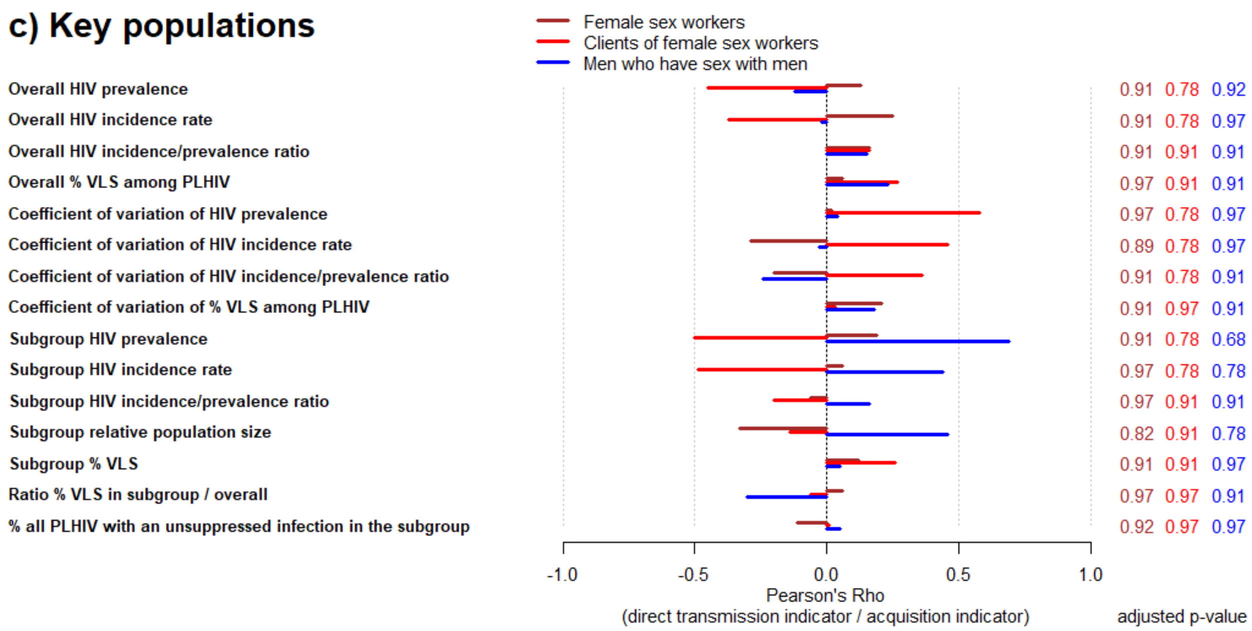

Figure S2: Univariate correlations between key model outcomes for 2020 and model estimated population-specific ratios of the *direct transmission* and *acquisition* indicators for 2020 ($\frac{I_2(2020)}{I_1(2020)}$) among a) non-key population women, b) non-key population men, and c) key populations, using Pearson's correlation tests. Test p-values were adjusted using the Benjamini-Hochberg procedure. Younger: aged 15-24 years old; Older: aged 25+ years old.

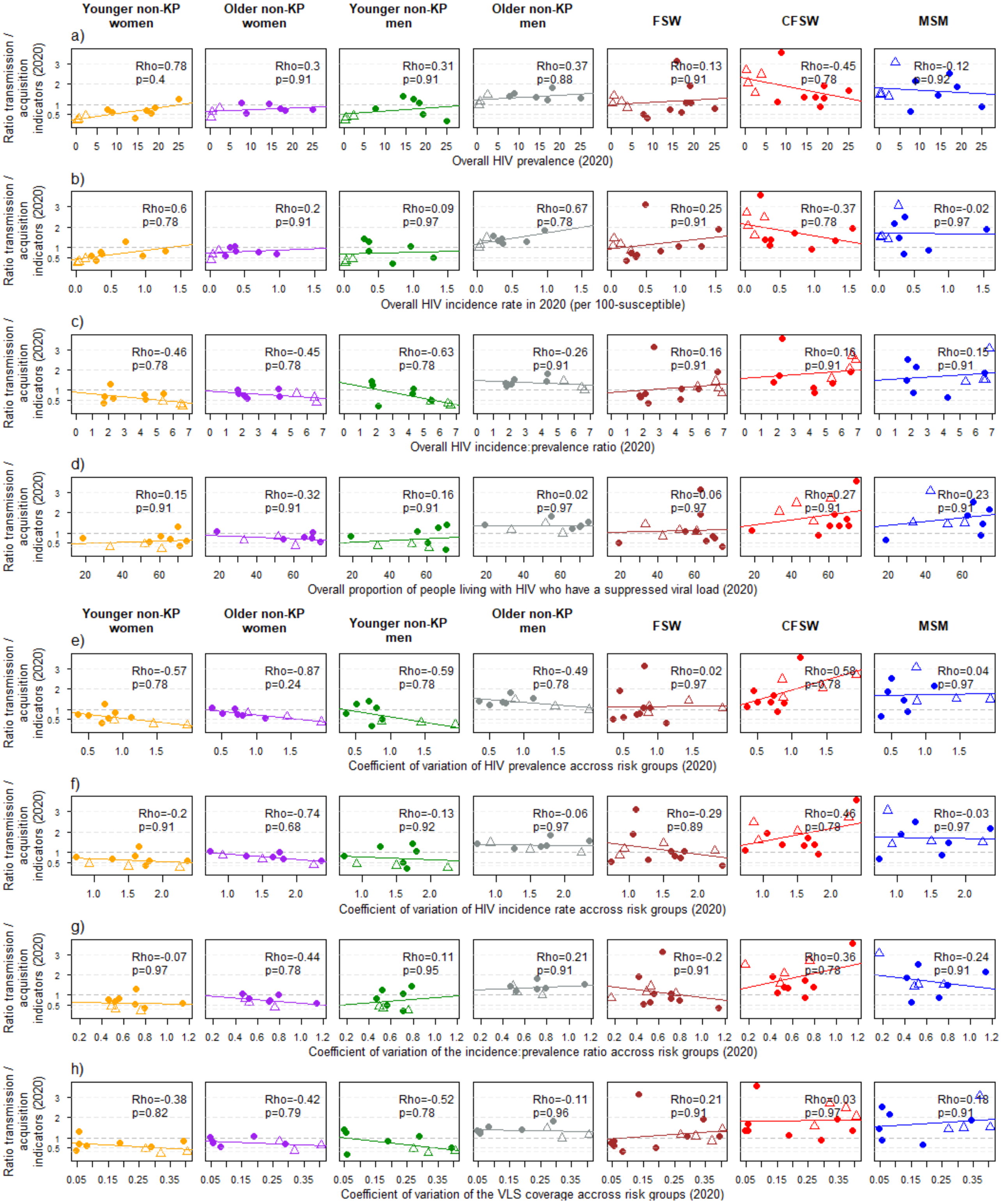

Younger non-KP women
Older non-KP women
Younger non-KP men
Older non-KP men
FSW
CFSW
MSM
Ratio transmission / acquisition indicators (2020)
a)
Rho=0.78 p=0.4
Rho=0.3 p=0.91
Rho=0.31 p=0.91
Rho=0.37 p=0.88
Rho=0.13 p=0.91
Rho=-0.45 p=0.78
Rho=-0.12 p=0.92
Overall HIV prevalence (2020)
b)
Rho=0.6 p=0.78
Rho=0.2 p=0.91
Rho=0.09 p=0.97
Rho=0.67 p=0.78
Rho=0.25 p=0.91
Rho=-0.37 p=0.78
Rho=-0.02 p=0.97
Overall HIV incidence rate in 2020 (per 100-susceptible)
c)
Rho=-0.46 p=0.78
Rho=-0.45 p=0.78
Rho=-0.63 p=0.78
Rho=-0.26 p=0.91
Rho=0.16 p=0.91
Rho=0.16 p=0.91
Rho=0.15 p=0.91
Overall HIV incidence:prevalence ratio (2020)
d)
Rho=0.15 p=0.91
Rho=-0.32 p=0.91
Rho=0.16 p=0.91
Rho=0.02 p=0.97
Rho=0.06 p=0.97
Rho=0.27 p=0.91
Rho=0.23 p=0.91
Overall proportion of people living with HIV who have a suppressed viral load (2020)
Younger non-KP women
Older non-KP women
Younger non-KP men
Older non-KP men
FSW
CFSW
MSM
e)
Rho=-0.57 p=0.78
Rho=-0.87 p=0.24
Rho=-0.59 p=0.78
Rho=-0.49 p=0.78
Rho=0.02 p=0.97
Rho=0.59 p=0.78
Rho=0.04 p=0.97
Coefficient of variation of HIV prevalence accross risk groups (2020)
f)
Rho=-0.2 p=0.91
Rho=-0.74 p=0.68
Rho=-0.13 p=0.92
Rho=-0.06 p=0.97
Rho=-0.29 p=0.89
Rho=0.46 p=0.78
Rho=-0.03 p=0.97
Coefficient of variation of HIV incidence rate accross risk groups (2020)
g)
Rho=-0.07 p=0.97
Rho=-0.44 p=0.78
Rho=0.11 p=0.95
Rho=0.21 p=0.91
Rho=-0.2 p=0.91
Rho=0.36 p=0.78
Rho=-0.24 p=0.91
Coefficient of variation of the incidence:prevalence ratio accross risk groups (2020)
h)
Rho=-0.38 p=0.82
Rho=-0.42 p=0.79
Rho=-0.52 p=0.78
Rho=-0.11 p=0.96
Rho=0.21 p=0.91
Rho=0.03 p=0.97
Rho=0.48 p=0.91
Coefficient of variation of the VLS coverage accross risk groups (2020)

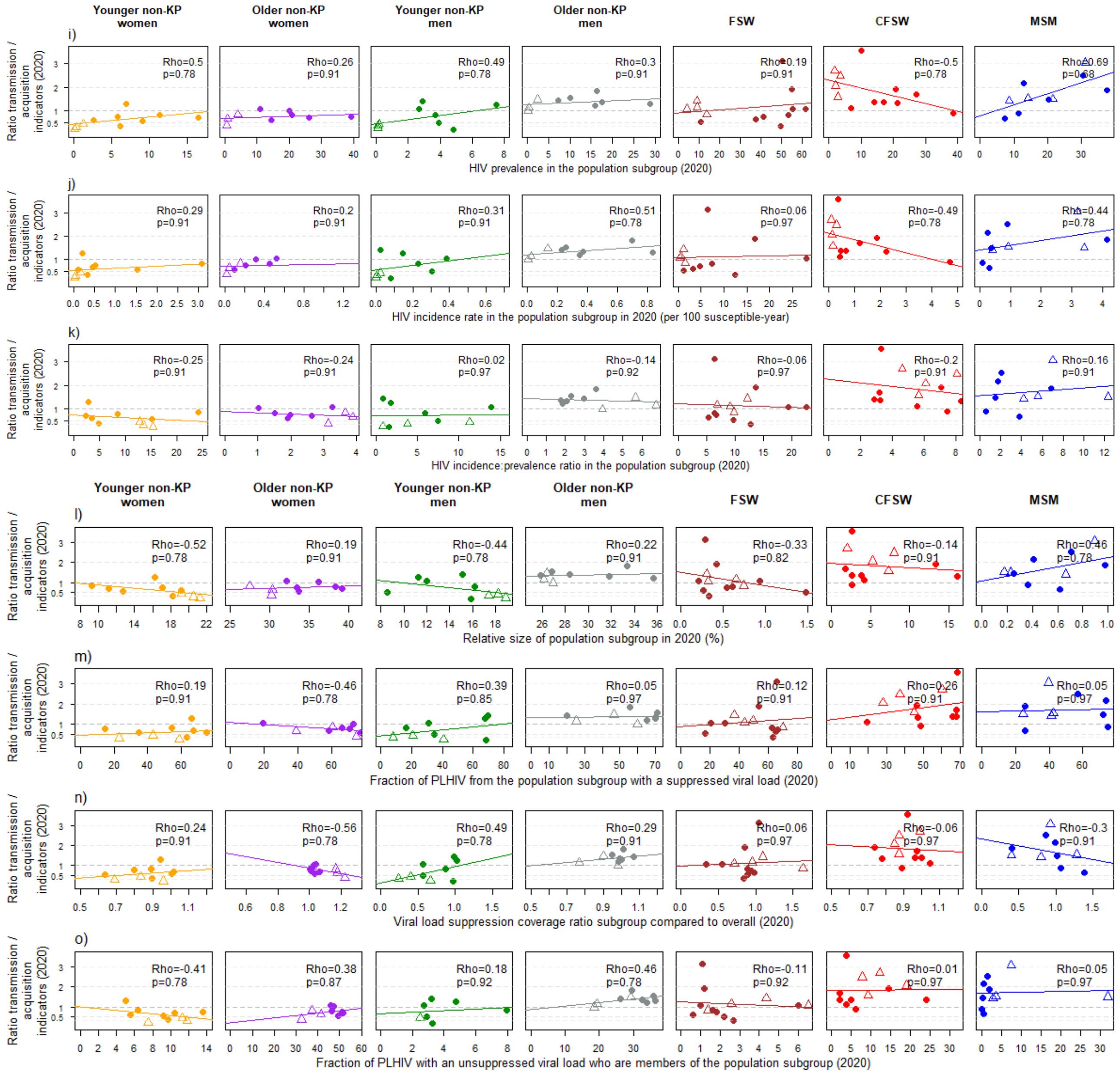

Figure S3: Correlation between model estimated population-specific ratios of the *direct transmission* and *acquisition* indicators for 2020 ($\frac{I_2(2020)}{I_1(2020)}$, y-axis), and the overall a) HIV prevalence, b) HIV incidence rate (expressed per 100 susceptible-year), c) HIV incidence/prevalence ratio, d) fraction of people living with HIV who have a suppressed viral load, and the coefficients of variation (calculated as the standard deviation of the output across populations over the average output across populations) of the overall e) HIV prevalence, f) HIV incidence rate (expressed per 100 susceptible-year), g) HIV incidence/prevalence ratio, and h) fraction of people living with HIV who have a suppressed viral load, the population-specific i) HIV prevalence, j) HIV incidence rate (expressed per 100 susceptible-year), and k) HIV incidence/prevalence ratio, l) relative size of the population (relative to all adults aged 15+ years old), m) fraction of people living with HIV who have a suppressed HIV viral load, n) ratio of the coverage of HIV viral suppression compared to overall, and o) fraction of PLHIV with an unsuppressed viral load who are members of the population (i.e., distribution of unsuppressed HIV), calculated in 2020 for each population in Eastern and Southern Africa (dots) and Western and Central Africa (triangles). Solid coloured lines are regression lines from a linear model. Each panel report estimates of Spearman's correlation coefficient and adjusted p-values. To improve the readability of the figure, we have not displayed the estimates for younger non-KP males from the Optima model for Malawi (which were included in our correlation tests), with *acquisition* and *direct transmission indicators* for 2020 of 0.07% and 0.59%, respectively. Younger: aged 15-24 years old; Older: aged 25+ years old; FSW: female sex workers; CFSW: clients of female sex workers; MSM: men who have sex with men; KP: key populations (including female sex workers, their clients, and men who have sex with men)

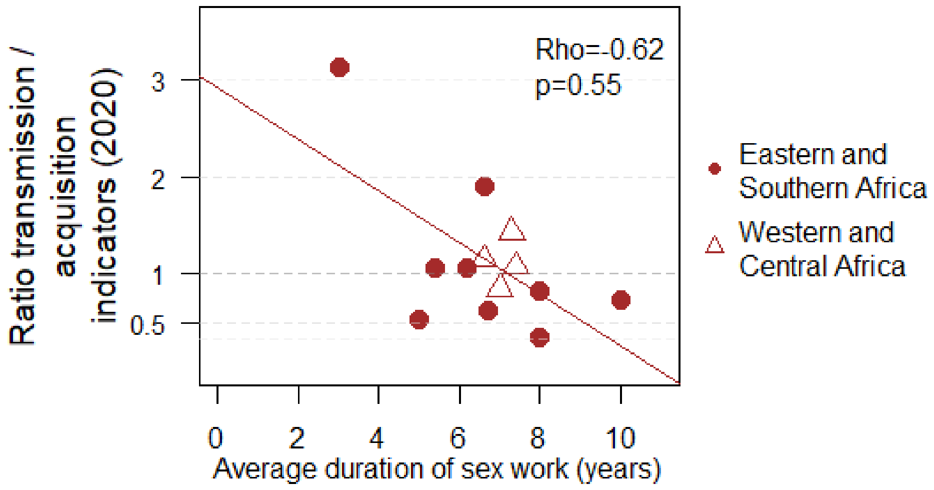

Figure S4: Correlation between the model average duration of sex work among female sex workers (x-axis) and the ratio of the *direct transmission* and *acquisition* indicators for 2020 ($\frac{I_2(2020)}{I_1(2020)}$, y-axis), with estimated Spearman's correlation coefficient and adjusted p-values.

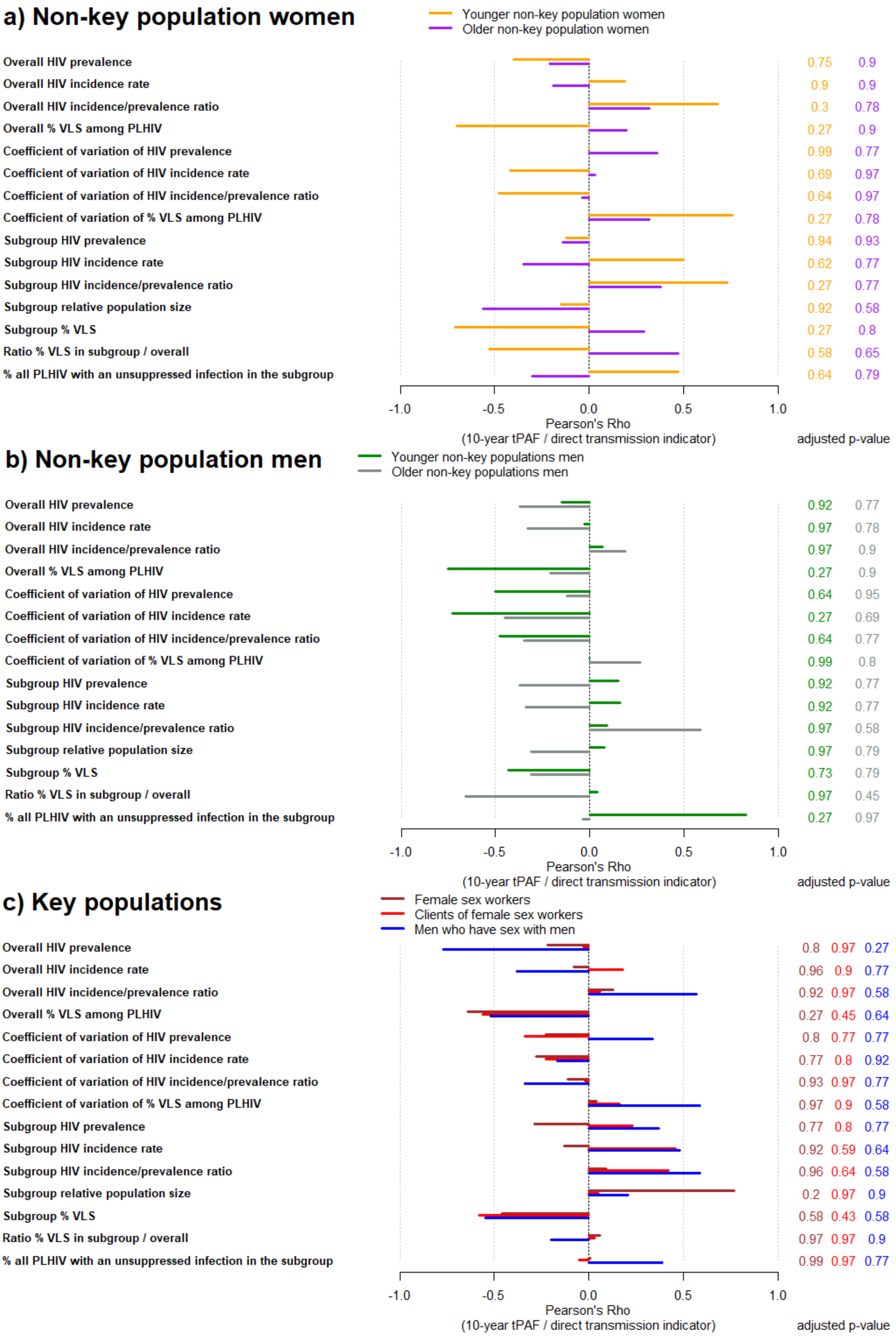
a) Non-key population women
Younger non-key population women
Older non-key population women
Overall HIV prevalence 0.75 0.9
Overall HIV incidence rate 0.9 0.9
Overall HIV incidence/prevalence ratio 0.3 0.78
Overall % VLS among PLHIV 0.27 0.9
Coefficient of variation of HIV prevalence 0.99 0.77
Coefficient of variation of HIV incidence rate 0.69 0.97
Coefficient of variation of HIV incidence/prevalence ratio 0.64 0.97
Coefficient of variation of % VLS among PLHIV 0.27 0.78
Subgroup HIV prevalence 0.94 0.93
Subgroup HIV incidence rate 0.62 0.77
Subgroup HIV incidence/prevalence ratio 0.27 0.77
Subgroup relative population size 0.92 0.58
Subgroup % VLS 0.27 0.8
Ratio % VLS in subgroup / overall 0.58 0.65
% all PLHIV with an unsuppressed infection in the subgroup 0.64 0.79
-1.0 -0.5 0.0 0.5 1.0
Pearson's Rho
(10-year tPAF / direct transmission indicator)
adjusted p-value
b) Non-key population men
Younger non-key populations men
Older non-key populations men
Overall HIV prevalence 0.92 0.77
Overall HIV incidence rate 0.97 0.78
Overall HIV incidence/prevalence ratio 0.97 0.9
Overall % VLS among PLHIV 0.27 0.9
Coefficient of variation of HIV prevalence 0.64 0.95
Coefficient of variation of HIV incidence rate 0.27 0.69
Coefficient of variation of HIV incidence/prevalence ratio 0.64 0.77
Coefficient of variation of % VLS among PLHIV 0.99 0.8
Subgroup HIV prevalence 0.92 0.77
Subgroup HIV incidence rate 0.92 0.77
Subgroup HIV incidence/prevalence ratio 0.97 0.58
Subgroup relative population size 0.97 0.79
Subgroup % VLS 0.73 0.79
Ratio % VLS in subgroup / overall 0.97 0.45
% all PLHIV with an unsuppressed infection in the subgroup 0.27 0.97
-1.0 -0.5 0.0 0.5 1.0
Pearson's Rho
(10-year tPAF / direct transmission indicator)
adjusted p-value
c) Key populations
Female sex workers
Clients of female sex workers
Men who have sex with men
Overall HIV prevalence 0.8 0.97 0.27
Overall HIV incidence rate 0.96 0.9 0.77
Overall HIV incidence/prevalence ratio 0.92 0.97 0.58
Overall % VLS among PLHIV 0.27 0.45 0.64
Coefficient of variation of HIV prevalence 0.8 0.77 0.77
Coefficient of variation of HIV incidence rate 0.77 0.8 0.92
Coefficient of variation of HIV incidence/prevalence ratio 0.93 0.97 0.77
Coefficient of variation of % VLS among PLHIV 0.97 0.9 0.58
Subgroup HIV prevalence 0.77 0.8 0.77
Subgroup HIV incidence rate 0.92 0.59 0.64
Subgroup HIV incidence/prevalence ratio 0.96 0.64 0.58
Subgroup relative population size 0.2 0.97 0.9
Subgroup % VLS 0.58 0.43 0.58
Ratio % VLS in subgroup / overall 0.97 0.97 0.9
% all PLHIV with an unsuppressed infection in the subgroup 0.99 0.97 0.77
-1.0 -0.5 0.0 0.5 1.0
Pearson's Rho
(10-year tPAF / direct transmission indicator)
adjusted p-value

Figure S5: Univariate correlations between key model outcomes for 2020 and model estimated population-specific ratios of the 10-year tPAF (2020-2029) over the *direct transmission indicator* for 2020 ($\frac{I_4(2020-2029)}{I_2(2020)}$) among a) non-key population women, b) non-key population men, and c) key populations, using Pearson's correlation tests. Test p-values were adjusted using the Benjamini-Hochberg procedure. Younger: aged 15-24 years old; Older: aged 25+ years old.

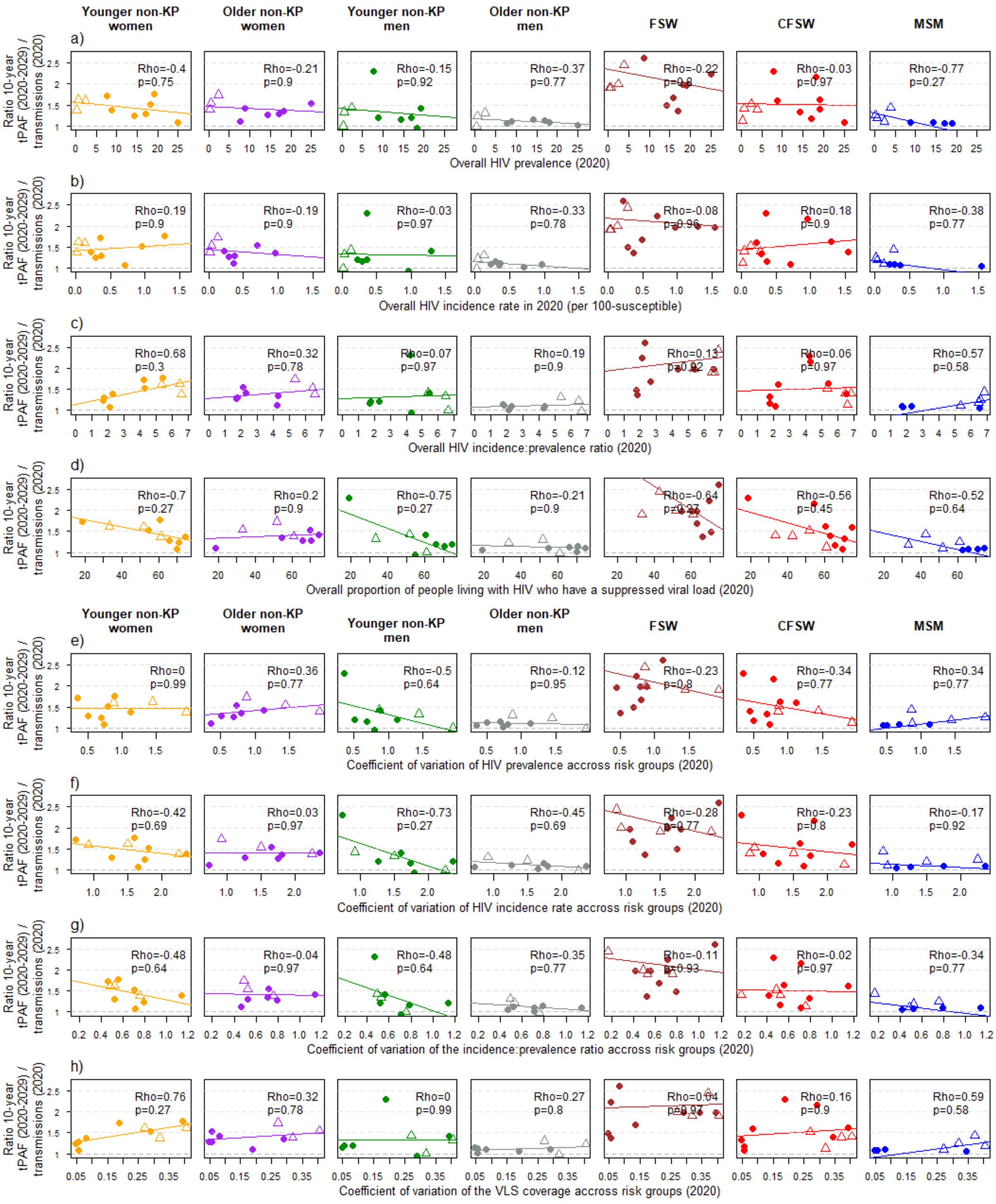

Younger non-KP women
Older non-KP women
Younger non-KP men
Older non-KP men
FSW
CFSW
MSM
Ratio 10-year tPAF (2020-2029) / transmissions (2020)
a)
Rho=-0.4 p=0.75
Rho=-0.21 p=0.9
Rho=-0.15 p=0.92
Rho=-0.37 p=0.77
Rho=-0.22 p=0.8
Rho=-0.03 p=0.97
Rho=-0.77 p=0.27
Overall HIV prevalence (2020)
b)
Rho=0.19 p=0.9
Rho=-0.19 p=0.9
Rho=-0.03 p=0.97
Rho=-0.33 p=0.78
Rho=-0.08 p=0.96
Rho=0.18 p=0.9
Rho=-0.38 p=0.77
Overall HIV incidence rate in 2020 (per 100-susceptible)
c)
Rho=0.68 p=0.3
Rho=0.32 p=0.78
Rho=0.07 p=0.97
Rho=0.19 p=0.9
Rho=0.13 p=0.92
Rho=0.06 p=0.97
Rho=0.57 p=0.58
Overall HIV incidence:prevalence ratio (2020)
d)
Rho=-0.7 p=0.27
Rho=0.2 p=0.9
Rho=-0.75 p=0.27
Rho=-0.21 p=0.9
Rho=-0.64 p=0.27
Rho=-0.56 p=0.45
Rho=-0.52 p=0.64
Overall proportion of people living with HIV who have a suppressed viral load (2020)
Younger non-KP women
Older non-KP women
Younger non-KP men
Older non-KP men
FSW
CFSW
MSM
e)
Rho=0 p=0.99
Rho=0.36 p=0.77
Rho=-0.5 p=0.64
Rho=-0.12 p=0.95
Rho=-0.23 p=0.8
Rho=-0.34 p=0.77
Rho=0.34 p=0.77
Coefficient of variation of HIV prevalence accross risk groups (2020)
f)
Rho=-0.42 p=0.69
Rho=0.03 p=0.97
Rho=-0.73 p=0.27
Rho=-0.45 p=0.69
Rho=-0.28 p=0.77
Rho=-0.23 p=0.8
Rho=-0.17 p=0.92
Coefficient of variation of HIV incidence rate accross risk groups (2020)
g)
Rho=-0.48 p=0.64
Rho=-0.04 p=0.97
Rho=-0.48 p=0.64
Rho=-0.35 p=0.77
Rho=-0.11 p=0.93
Rho=-0.02 p=0.97
Rho=-0.34 p=0.77
Coefficient of variation of the incidence:prevalence ratio accross risk groups (2020)
h)
Rho=0.76 p=0.27
Rho=0.32 p=0.78
Rho=0 p=0.99
Rho=0.27 p=0.8
Rho=0.04 p=0.97
Rho=0.16 p=0.9
Rho=0.59 p=0.58
Coefficient of variation of the VLS coverage accross risk groups (2020)

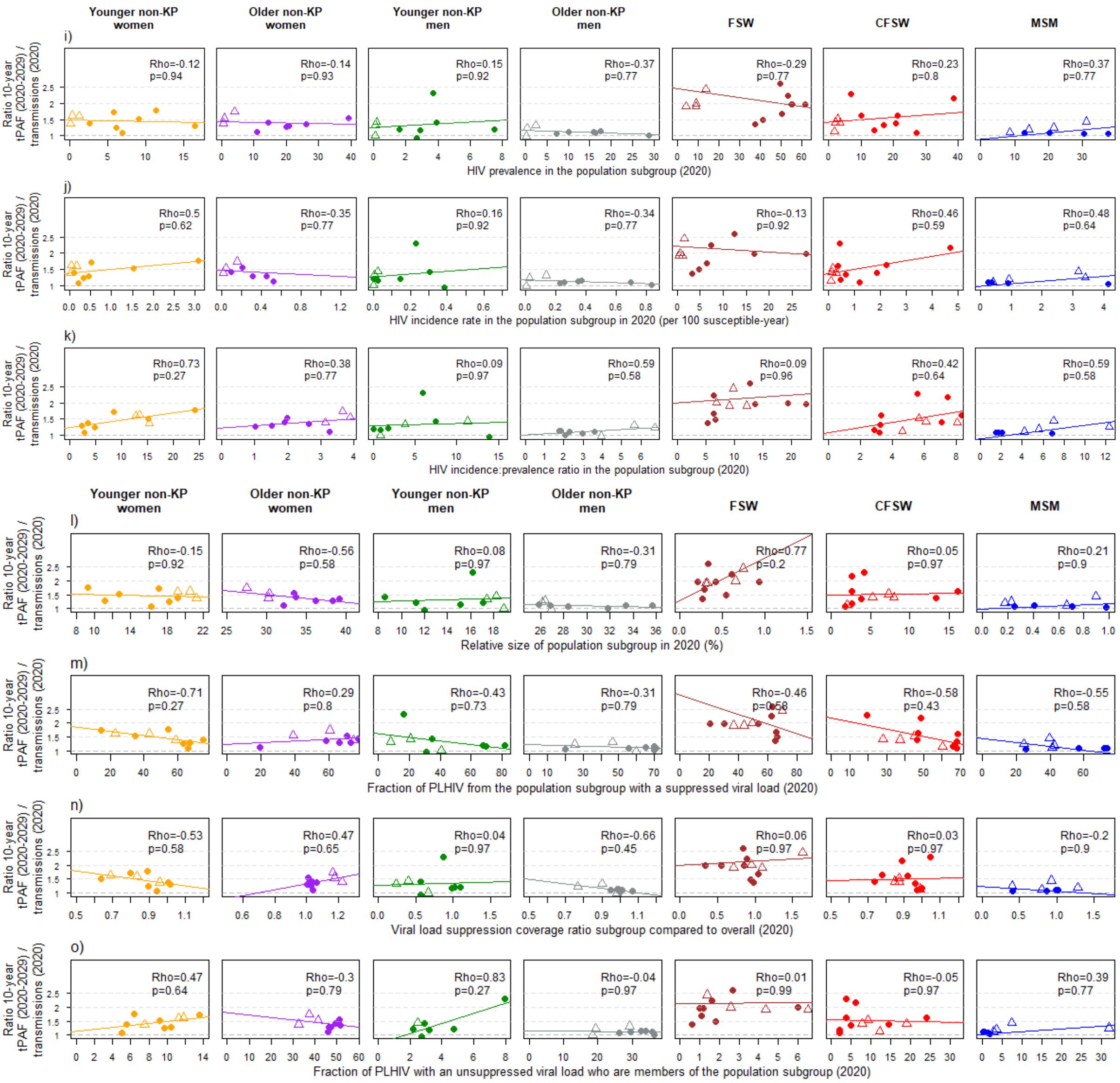

Figure S6: Correlation between model estimated population-specific ratios of the 10-year *tPAF* (2020-2029) and *direct transmission* indicators (2020) ($\frac{I_4(2020-2029)}{I_2(2020)}$, y-axis), and the overall a) HIV prevalence, b) HIV incidence rate (expressed per 100 susceptible-year), c) HIV incidence/prevalence ratio, d) fraction of people living with HIV who have a suppressed viral load, and the coefficients of variation (calculated as the standard deviation of the output across populations over the average output across populations) of the overall e) HIV prevalence, f) HIV incidence rate (expressed per 100 susceptible-year), g) HIV incidence/prevalence ratio, and h) fraction of people living with HIV who have a suppressed viral load, the population-specific i) HIV prevalence, j) HIV incidence rate (expressed per 100 susceptible-year), and k) HIV incidence/prevalence ratio, l) relative size of the population (relative to all adults aged 15+ years old), m) fraction of people living with HIV who have a suppressed HIV viral load, n) ratio of the coverage of HIV viral suppression compared to overall, and o) fraction of PLHIV with an unsuppressed viral load who are members of the population (i.e., distribution of unsuppressed HIV), calculated in 2020 for each population in Eastern and Southern Africa (dots) and Western and Central Africa (triangles). Solid coloured lines are regression lines from a linear model. Each panel report estimates of Spearman's correlation coefficient and adjusted p-values. To improve the readability of the figure, we have not displayed the estimates for younger non-KP males from the Optima model Eswatini (which were included in our correlation tests), with direct transmission indicators for 2020 and 10-year tPAFs over 2020-2029 of 0.47% and 2.22%, respectively, as well as estimates for MSM from the Optima model for Mozambique, with direct transmission indicators for 2020 and 10-year tPAFs over 2020-2029 of 0.34% and 5.06%, respectively. Younger: aged 15-24 years old; Older: aged 25+ years old; FSW: female sex workers; CFSW: clients of female sex workers; MSM: men who have sex with men; KP: key populations (including female sex workers, their clients, and men who have sex with men)

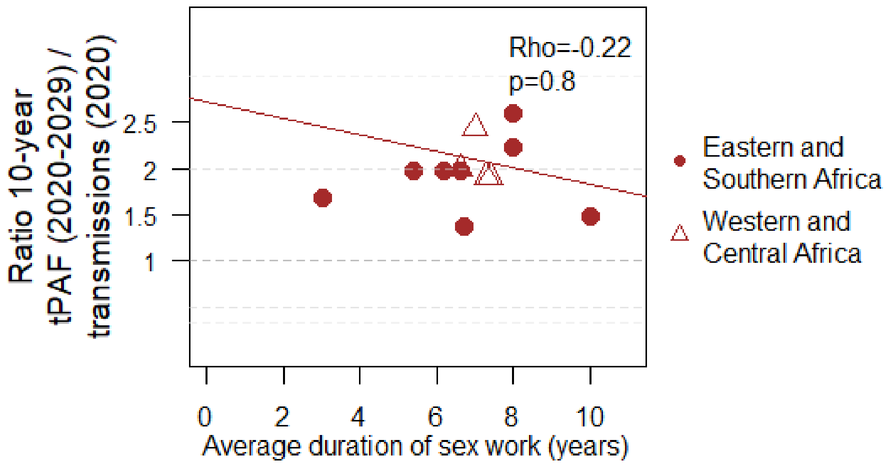

Figure S7: Correlation between the model average duration of sex work among female sex workers (x-axis) and the ratio between the 10-year tPAF (2020-2029) and the *direct transmission* indicator for 2020 ($\frac{I_4(2020-2029)}{I_2(2020)}$, y-axis), with estimated Spearman's correlation coefficient and adjusted p-values.

### Supplement results: largest contributors to new HIV infections in 2010 and 2010-2019

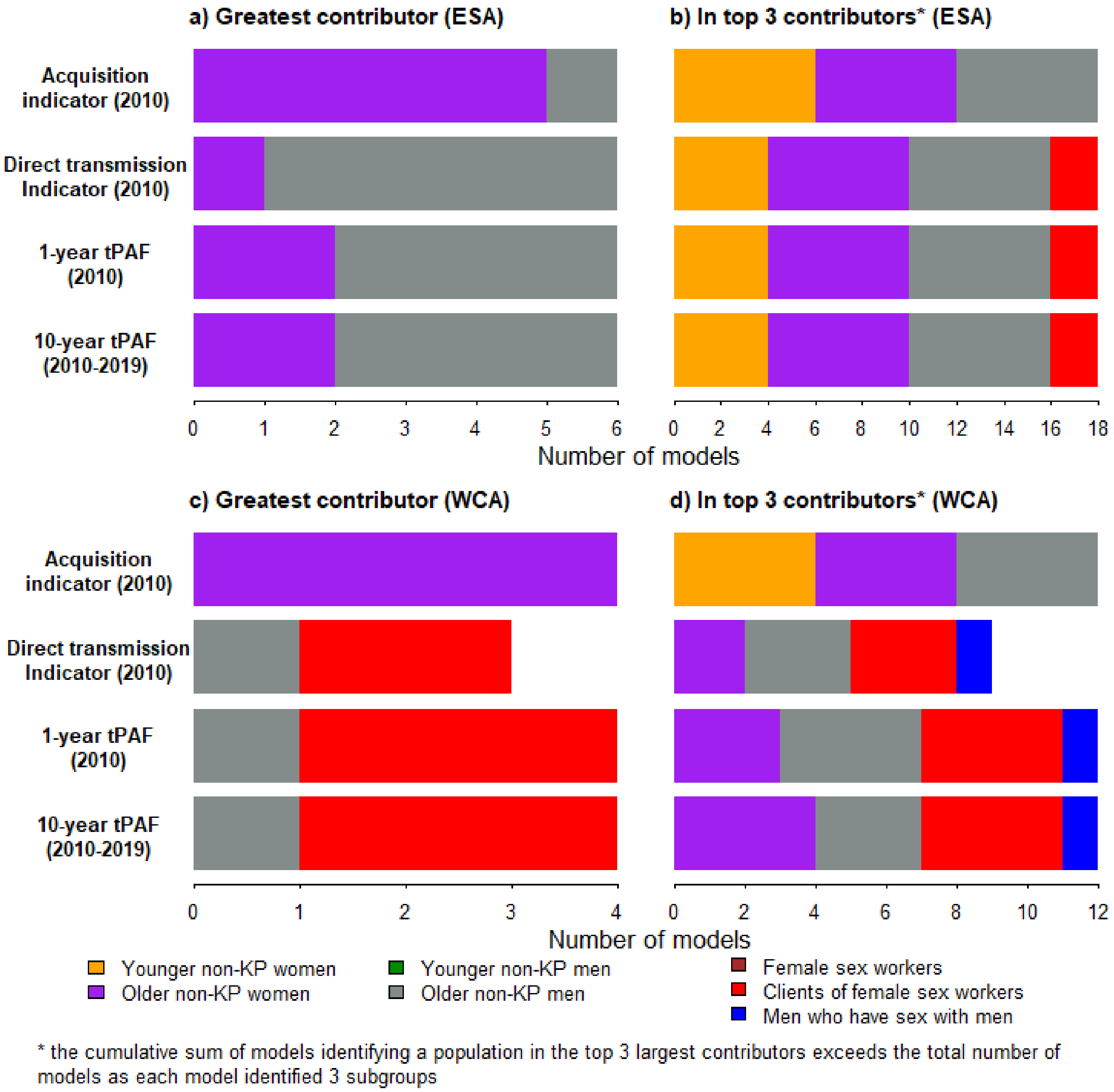

Figure S8. Differences in the populations identified as greatest (left panels) and among top 3 greatest (right panels) contributors to new HIV infections by indicators from mathematical models for a-b) Eastern and Southern Africa, and c-d) Western and Central Africa. $I_1$: acquisition indicator (2010), $I_2$: direct transmission indicator (2010), $I_3$: 1-year transmission population-attributable fraction (2010), and $I_4$: 10-year transmission population-attributable fraction (2010-2019). One model providing estimates for all selected populations in Western and Central Africa did not report estimates of the direct transmission indicator. None of the

10 models identified female sex workers and non-key population men among the top 3 largest contributors in any of the 4 indicators, whereas no model for Eastern and Southern Africa identifies men who have sex with men among the top 3 largest contributors in any of the 4 indicators. Additional details are provided in Table S2.

Younger: aged 15-24 years old; Older: aged 25+ years old; KP: key populations (including female sex workers, their clients, and men who have sex with men).

### Supplement results: comparison between indicator ratios over two periods

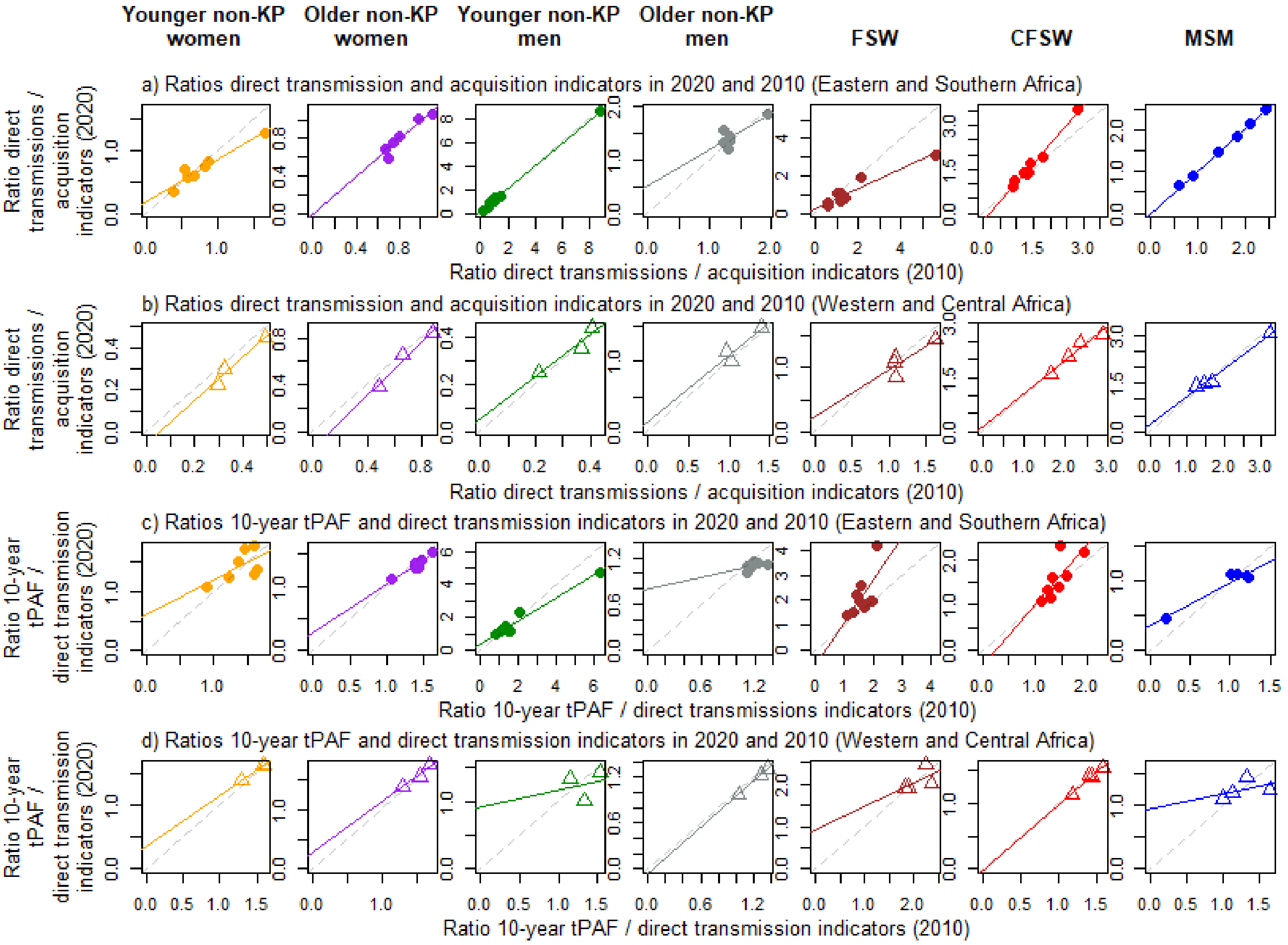

Figure S9. Ratios of the *direct transmission indicator* over the *acquisition indicator* ($\frac{I_2}{I_1}$) calculated over 2010 (x-axis) and 2020 (y-axis) in a) Eastern and Southern Africa, and b) Western and Central Africa. Ratios of the 10-year tPAF over the *direct transmission indicator* ($\frac{I_4}{I_2}$) calculated over different time points ($\frac{I_4(2010-2019)}{I_2(2010)}$ on the x-axis, and

$\frac{I_4(2020-2029)}{I_2(2020)}$ on the y-axis) in c) Eastern and Southern Africa, and d) Western and Central Africa. Grey dashed diagonal lines indicate perfect agreement between indicator ratios in 2010 and 2020, whereas colour plain lines are regression lines from a linear model.

Younger: aged 15-24 years old; Older: aged 25+ years old; FSW: female sex workers; CFSW: clients of female sex workers; MSM: men who have sex with men; KP: key populations (including female sex workers, their clients, and men who have sex with men)
